# Variation in menstrual cycle length by age, race/ethnicity, and body mass index in a large digital cohort of women in the US

**DOI:** 10.1101/2022.09.30.22280382

**Authors:** Huichu Li, Elizabeth A. Gibson, Anne Marie Z. Jukic, Donna D. Baird, Allen J. Wilcox, Christine L. Curry, Tyler Fischer-Colbrie, Jukka-Pekka Onnela, Michelle A. Williams, Russ Hauser, Brent A. Coull, Shruthi Mahaligaiah

## Abstract

**Background:** Menstrual characteristics are important signs of women’s health. We examined the variation of menstrual cycle length by age, race and ethnicity, and body weight using data collected from mobile menstrual tracking apps. Understanding how menstrual characteristics vary by these factors can provide important information for further study of environmental and social determinants of menstrual health.

**Methods:** We collected self-tracked menstrual cycle data from participants of the Apple Women’s Health Study. Demographic and lifestyle characteristics were self-reported from surveys. Linear mixed effect (LME) models were used to estimate the differences in cycle length associated with age, race/ethnicity, and body mass index (BMI), adjusted for possible confounders or predictors of cycle length. Cycle variability was estimated by the change of within-individual standard deviations of cycle length.

**Findings:** A total of 165,668 cycles from 12,608 participants from US were included. After adjusting for all covariates, mean menstrual cycle length was shorter with older age across all age groups until age 50 and then became longer for those age 50 and older. Menstrual cycles were on average 1·6 (95%CI: 1·2, 2·0) days longer for Asian and 0·7 (0·4, 1·0) days longer for Hispanic participants compared to White non-Hispanic participants. Participants with Class 3 obesity (BMI≥40 kg/m^2^) had 1·5 (1·2, 1·8) days longer cycles compared to those with healthy BMI (18·5≤BMI<25 kg/m^2^). Cycle variability was smaller among participants in older age groups but became considerably larger for those in the oldest age categories (45-49 and 50+). Asian and Hispanic participants and those who were obese had larger cycle variability.

**Interpretation:** This study demonstrated differences in menstrual characteristics by age, race and ethnicity, and obesity using data collected from mobile health apps. Future studies should explore the underlying determinants of the racial and ethnic differences in menstrual characteristics.

**Funding:** Apple Inc.

**Research in context:** *Evidence before this study:* We searched PubMed for studies on menstrual cycles with age, body weight, and race/ethnicity using the term “((“Menstrual Cycle”[Mesh:NoExp] OR menstrual cycle*[tiab]) AND (“Time Factors”[Mesh] OR cycle length*[tiab] OR variability[tiab] OR variation*[tiab])) OR (“Menstruation”[Mesh] AND (“Time Factors”[Mesh] OR length[tiab] OR variability[tiab] OR variation*[tiab]) AND 1950[pdat]:1986[pdat]) AND (“Age Factors”[Mesh:NoExp] OR “Race Factors”[Mesh] OR “Racial Groups”[Mesh] OR “Ethnicity”[Mesh] OR “ethnology” [Subheading] OR “Body Mass Index”[Mesh] OR “Body Weight”[Mesh:NoExp] OR “Overweight”[Mesh] OR age[tiab] OR ages[tiab] OR race[tiab] OR races[tiab] OR racial[tiab] OR ethnic*[tiab] OR body mass[tiab] OR bmi[tiab] OR weight[tiab] OR overweight[tiab] OR obes*[tiab]) NOT (“Animals”[Mesh] NOT “Humans”[Mesh])” from 1950 to August 3, 2022. This search yielded 2,064 sources and we identified 30 research articles comparing menstrual cycle length and/or variability by age, race/ethnicity, and/or body weight in population. Another 14 research articles were added to the search results by knowledge of existing literature. Of the 44 articles identified, studies on changes of menstrual cycle length and variability with age reported consistent results, although the age range of the participants differed across these studies. In general, menstrual cycles were longer and less regular in the first few years following menarche, but over the subsequent decades cycles tend to become shorter and more regular with older age until age 40-45, after which they can become increasingly longer and irregular until menopause. Obesity was associated with menstrual irregularity. However, the association with menstrual cycle length was less consistently reported. Some studies found obesity was associated with longer menstrual cycles while other studies reported null associations. Compared to age and body weight, fewer studies have considered racial and ethnic differences of menstrual characteristics, although results from separate studies in Japan, China, and India suggested that females in these countries had longer cycle lengths compared to those observed in White females in US. Many of the prior studies of menstrual cycle characteristics relied on self-reported typical menstrual cycle length and regularity in survey questions, without acquiring actual menstrual cycle data. Most of those that used menstrual diaries to obtain cycle characteristics with higher accuracy were limited by relatively small numbers of participants. A few recent studies using menstrual tracking app data from a large number of users in different countries (mainly in Europe and North America) reported similar associations of age with cycle length and variability. However, results for body weight were still inconsistent. In addition, menstrual cycle characteristics by race and ethnicity have seldom been characterized using such data.

*Added value of this study:* In this large digital cohort study, we collected menstrual cycle data from menstrual tracking apps and factors related to menstruation from surveys to comprehensively compare the distribution of menstrual cycle length by age, race and ethnicity, and body mass index in a diverse US population. Using this dataset, our study results confirmed the non-linear changes of menstrual pattern with age throughout the reproductive lifespan as characterized in previous studies. More importantly, after controlling for potential confounders, we observed racial and ethnic differences of menstrual cycle length, in which participants who were Asian and who were Hispanic had longer menstrual cycles and higher variability. Obesity was associated with longer menstrual cycle length and higher cycle variability, especially among Hispanic individuals.

*Implications of all the available evidence:* Menstrual cycle characteristics such as cycle length and variability/regularity have been recognized as important vital signs associated with gynecological conditions, fertility, cardiometabolic diseases, and mortality risk. Previous studies provided different estimates on menstrual parameters and current recommendations on normal menstrual cycle length and variability were based on limited quantitative evidence that was mainly generated in the White population. With the popularity of menstrual cycle tracking apps, our study demonstrated that this app-based data is a promising and powerful resource for research of menstrual health today. This study also provided valuable evidence on possible racial and ethnic disparities in menstrual cycle characteristics in a large free-living population. Future studies should consider the underlying environmental, social, and behavioral factors that drive the observed racial and ethnic differences of menstrual characteristics.

## INTRODUCTION

Menstrual cycle characteristics, including cycle length and regularity (i.e., the variability of cycle length within an individual), have been recognized to be an important vital sign (1). Accumulating evidence has also documented associations of long and/or irregular menstrual cycles with higher risk of infertility, cardiometabolic disease, and death (2–6).

It has been shown that menstrual cycle length varies considerably within an individual throughout the reproductive lifespan (7–9). Several studies using menstrual diary data from small numbers of individuals reported decreasing length and variability of menstrual cycle with increasing age from late adolescence/early adulthood until late reproductive age (i.e., age 40-45) (9–14). Reports on menstrual characteristics were limited for women above age 45, and current evidence suggested menstrual cycles were increasingly longer and more varied in this age group (8,15,16). Obesity has been linked with longer and less regular menstrual cycles, however, the results were not consistent (10,14,16,17). The recent emergence of menstrual cycle tracking applications (apps) in smart phones allows large epidemiologic studies to confirm previous findings in small population samples and to examine factors of menstrual health in a rapidly changing world (18–20). Two studies using the app data reported similar changes of menstrual patterns by age as observed previously. However, results for body mass index (BMI) were still inconsistent (21,22). In addition, adjustment for confounding, such as age, race and ethnicity, diet, and physical activity, was limited in these studies.

Earlier evidence on menstrual characteristics was mainly from studies among White women and has been used to establish the normal range of menstrual cycle length for clinical practice (23,24). Separate observations among individuals in Japan, China, and India reported approximately 1-2 days longer cycle lengths compared to those observed in the White population, indicating a possibility that the suggested cycle pattern parameters may not be applicable in individuals with different racial and ethnic backgrounds (25–27). Other studies also found racial and ethnic differences in hormones related to menstruation (28–31). However, studies directly examining racial and ethnic differences in menstrual patterns were limited and included small groups of individuals (11,14,16,32). Therefore, evidence from a larger population is warranted.

In this study, we used menstrual cycle tracking data and survey information to confirm the associations between age and BMI with menstrual cycle length and cycle variability, and to examine possible differences of menstrual characteristics by race/ethnicity in a nationwide digital cohort of women within the United States (US). Our hypotheses are: there are non-linear relationships between age and each of menstrual cycle length and variation. Cycles are longer and more varied in the first few years after menarche, then become shorter and more regular with increasing age until menopausal transition, when cycles start to elongate and vary in length. Higher BMI is associated with longer length and higher variability in menstrual cycles. In addition, menstrual cycle characteristics differ across race and ethnicity groups.

## METHODS

We used the STROBE reporting guideline for this section.

### Study design and population

The Apple Women’s Health Study (AWHS) is an ongoing, prospective digital cohort study. Users of the Apple Research app on their iPhone were eligible if they have ever menstruated at least once in life, live in the US, were at least 18 years old (at least 19 in Alabama and Nebraska, and 21 in Puerto Rico), and are able to communicate in English. Eligibility also required sole usage of their iCloud account or iPhone. Enrollment began on November 2019 and is ongoing. Informed consent is provided at enrollment. This study has been approved by the Institutional Review Board at Advarra (CIRB #PRO00037562) and has been registered in Clinicaltrials.gov (NCT04196595).

Detailed information on the study design and data collection has been published (33). Briefly, participants are asked to complete surveys on demographic characteristics (e.g., race and ethnicity, height and weight, and socioeconomic status) and reproductive history. They are also asked to self-report gynecological conditions (e.g., polycystic ovarian syndrome (PCOS), uterine fibroids, and hysterectomy) and health behaviors (e.g., smoking, alcohol use, and physical activity) which are surveyed every 12 months during follow-up. Factors related to menstrual cycles, including hormone use, pregnancy, lactation, and menopause, are collected at enrollment and updated monthly in surveys. Information on cycle tracking accuracy was also collected in monthly surveys after enrollment.

For this analysis, we included eligible AWHS participants who did not report menopause, who enrolled and contributed at least one completed menstrual cycle by December 31, 2021, and who had no history of PCOS, uterine fibroids, or hysterectomy. We excluded participants with uterine fibroids because they may be more likely to experience intermenstrual bleeding, which may affect the accuracy of menstrual cycle identification (24).

### Menstrual cycle identification

Participants can track their menstrual flow using the Cycle Tracking feature with the Apple Health app or other third-party apps that the participant allows to write to the Health app. We collected menstrual flow entries prospectively after enrollment and any entries up to 24 months prior to enrollment. Spotting, defined as any bleeding that happens outside of the regular period, is not included in this analysis. A menstrual cycle was defined as one or more consecutive days with tracked menstrual flow followed by at least 2 days of no tracked flow. The first day of the having menstrual flow was identified as the first day of the cycle, as defined previously (8). Cycles shorter than 10 days or longer than 90 days are excluded from the analysis as they were unlikely for a natural menstrual cycle (21). For the remaining cycles, we excluded cycles that were atypically long and likely artifacts due to gaps in record-keeping using participant-specific thresholds modified from a previous study with details in the Supplemental Methods (19).

For menstrual cycles tracked after enrollment, we only included those that have been confirmed with no hormone use, pregnancy, or lactation in the monthly surveys. For cycles tracked prior to enrollment, we only included those from participants who confirmed none of these events in the previous 2 years.

### Measurement of age, race/ethnicity, and BMI

Age was calculated as the difference between the year of the first day of the cycle and the participant’s birth year and was categorized as under 20, 20-24, 25-29, 30-34, 35-39, 40-44, 45-49, and above 50 years. Race and ethnicity was self-reported using the following categories with instructions to check all that apply: White, non-Hispanic (referred to as ‘White’); Black or African American or African (referred to as ‘Black’); Asian; Hispanic, Latino, Spanish and/or other Hispanic (referred to as ‘Hispanic’); American Indian or Alaska Native; Middle Eastern or North African; Native Hawaiian or Pacific Islander; and an option indicating that none of these categories can fully describe the participant. For this analysis, we combined participants who were American Indian or Alaska Native, Middle Eastern or North African, Native Hawaiian or Pacific Islander, or who indicated none of these categories can fully describe the participant into one group because of small numbers. Participants who chose more than one category were combined in a separate group. Body mass index (BMI) was calculated using the self-reported height and weight. This was categorized as underweight (BMI<18·5 kg/m^2^), healthy (18·5≤BMI<25 kg/m^2^), overweight (25≤BMI<30 kg/m^2^), and obese (BMI≥30 kg/m^2^). The obese group was further divided into Class 1 (30≤BMI<35 kg/m^2^), 2 (35≤BMI<40 kg/m^2^), and 3 (BMI≥40 kg/m^2^) (34).

### Measurement of other covariates

We considered possible confounders or predictors of menstrual cycle length, including cigarette smoking (never smoked, previously smoked, and currently smoke), alcohol use, physical activity, stress, socioeconomic status, and parity (nulliparous and parous). All covariates were self-reported through surveys. Alcohol use was measured as frequency of up to once a month, 2-4 times a month, 2-3 times a week, and more than 4 times a week. Physical activity was categorized as none, light (e.g., walking or light housework), moderate (e.g., brisk walking or yard work), vigorous (e.g., running or carrying heavy loads), and strenuous (e.g., competitive sports or endurance events like marathons). Stress was measured using the 4-item Perceived Stress Score and categorized by quartiles (35). Socioeconomic status (SES) was measured using an objective and a subjective measure because it has been suggested that both measures could affect overall health jointly and independently (36). Highest education level was used as an objective measure of SES. The variable included levels: high school graduate or less (in the U.S. high school is grades 9-12), 3-year college or technical schools (typically done after graduating high school, not required for entrance to a 4-year college), 4-year college degree (typically begun directly after high school), and graduate school (including master and doctoral studies; occurs after receiving an undergraduate degree). The subjective measure of SES was measured by the MacArthur scale of subjective social status. This scale is a self-rated rank on a ‘social ladder’ from 0 (lowest) to 9 (highest) based on the responder’s self-perceived education, socioeconomic status, and current life circumstances relative to others. For this analysis, we categorized this scale into low (0-3), moderate (4-6), and high (7-9).

Each participant’s tracked menstrual cycles were merged with covariate values collected from the most recent surveys prior to that cycle. Menstrual cycles tracked prior to enrollment were assigned covariate values that were reported in their first surveys, corresponding to the assumption that these covariates remained unchanged during this interval. Participants with missing information on age, race/ethnicity, and BMI were excluded. Missingness in the other covariates was treated with missing indicators.

### Statistical analysis

We estimated the distribution of menstrual cycle length in our study population using a Gaussian kernel density function with weights equal to the inverse of the total number of cycles contributed by that participant to avoid bias by the varying numbers of menstrual cycles per participant.

#### Analysis of menstrual cycle length

We used linear mixed effect (LME) models with random participant-specific intercepts to estimate the differences and 95% confidence intervals (95%CIs) in mean menstrual cycle length by age, race/ethnicity, and BMI. To explore the possible influence of the other factors and covariates, we fitted a minimal model adjusted for age (univariable model when age is the main factor of interest) and a full model with age, race/ethnicity, BMI, and all other covariates included. We fitted a linear mixed quantile regression model for the median cycle length with age, race/ethnicity, and BMI, adjusted for all other covariates to examine the impact of cycle length distribution on the LME estimates (37). To better understand how the distribution of cycle length varies with these factors, we additionally considered the 25^th^ (P25) and 75^th^ percentiles (P75) of cycle length in the mixed quantile regression model. Pair-wise effect modification of age, race/ethnicity, and BMI was considered by adding multiplicative terms in the models. We restricted the effect modification analysis to participants who were under age 50 years and had BMI<40 kg/m^2^, and to White, Black, Asian, and Hispanic participants to avoid having strata with few cycles (Table S1). We then categorized menstrual cycles into long (>38 days) and short (<24 days) cycles using recommendations from the International Federation of Gynecology and Obstetrics (FIGO) (23), and examined the associations of each primary characteristic with the probability of experiencing a long or short menstrual cycle using logistic regression. The reference category for long and short cycles is menstrual cycles between 24-38 days. Participants with short cycles can contribute more cycles than those with long cycles, resulting in biased estimates from generalized estimating equations (due to informative cluster size). To address this bias, we estimated the odds ratio (OR) and 95%CI of experiencing a long or short cycle from logistic regression using the within-cluster resampling approach, adjusting for all covariates (Supplemental Methods) (38).

Several sensitivity analyses were considered. We repeated the analysis among participants who contributed at least three menstrual cycles because the individual-specific threshold may not effectively identify artifacts if a participant contributed very few cycles. Information on cycle tracking accuracy and most covariates was only available after enrollment. Therefore, we repeated the analysis by restricting to cycles tracked after enrollment with confirmed accuracy to examine the impact of possible measurement errors. An accurately tracked cycle was defined as a cycle started in a month when the participant responded ‘yes, they were accurate’ to the question ‘are all your period days during the previous calendar month accurately reflected in the Health app?’ in the corresponding monthly survey. We considered a complete case analysis for possible bias from missing indicators. We also repeated the analysis by including participants with uterine fibroids (N=705 participants). Since part of our data was collected during the COVID-19 pandemic, we considered a sensitivity analysis restricting to those who reported never having had a known COVID-19 infection in a 2022 survey.

#### Analysis of cycle length variability and irregularity

Considering women with few cycles may not effectively contribute information on cycle variability, we restricted this analysis to participants with at least three menstrual cycles. In the linear mixed model framework, within-individual variability in cycle length, which represents the degree of cycle irregularity, can be estimated by the standard deviations (SDs) of the model residuals after accounting for the systematic variability across subgroups in the fixed effect terms and between individual variability in random intercepts (8). To quantify and examine the associations of age, race/ethnicity, and BMI with within-individual cycle variability, we constructed log-linear models for residual variance in the fully adjusted LME models. We first considered univariable models to estimate within-individual cycle variability (in days) by each factor. Then we fitted a multivariable model with all three variables included to obtain the adjusted estimates for the associations of age, race/ethnicity, and BMI with within-individual cycle variability. Coefficients from the multivariable model were computed as the percentage change in within-individual variability in cycle length compared to the referent for the factors of interest.

We further identified cycle irregularity as participants whose mean difference in lengths of adjacent menstrual cycles ≥7 days (39) and examined the associations of age, race/ethnicity, and BMI with cycle irregularity using logistic regression models with iterative reweighted least squares, adjusted for all covariates. We repeated the analysis using alternative criteria of irregularity as the median difference in lengths of adjacent menstrual cycles ≥ 9 days (19), the standard deviation of menstrual cycle length ≥7 days, and the difference between the shortest and longest cycle ≥7 days.

Data management, processing, and statistical analyses were conducted in R (version 3·6·0) and Python (version 3·6). All statistical tests were two-sided. Data were accessed through servers hosted by Apple. While all authors on this manuscript are offered access to the study data, not all authors have accessed the data. HL and EAG (both not employed by Apple) have accessed and verified the data.

#### Role of Funding Source

The funding source provided platforms and tools for data collection and participated in writing of the manuscript. It played no role in the analysis and interpretation of data, and in the decision to submit.

## Results

A total of 794,282 menstrual cycles from 52,117 participants enrolled in the AWHS by December 31, 2021 were initially identified. After applying the exclusion criteria, a total of 165,668 menstrual cycles from 12,608 participants were included in the final analysis with a median of 11 cycles per participant (interquartile range, IQR = 5, 20) (Figure S1). A total of 64,326 cycles were tracked after enrollment, and among them 85% (N=54,804) were confirmed to be accurate. Approximately 88% (N=11,040) of participants had at least three menstrual cycles. Mean age of eligible participants at baseline was 33 years old (SD = 8) and over 70% of the participants were White. Nearly 35% (N=4,379) of the participants were obese (Table 1). The distribution of menstrual cycle length peaked at 28 days and had a long right tail (Figure S2, Table S2). The mean (SD) of this distribution was 28·7 days (6·1). The median (IQR) was 28 days (26, 30 days) and the 5^-^95^th^ percentile was 22-38 days. A total of 8,153 (5%) long cycles and 14,976 (9%) short cycles were identified. A total of 5,683 participants reported their history of COVID-19 infection, among them 4,119 reported never had known COVID-19 infection.

**Table 1.**
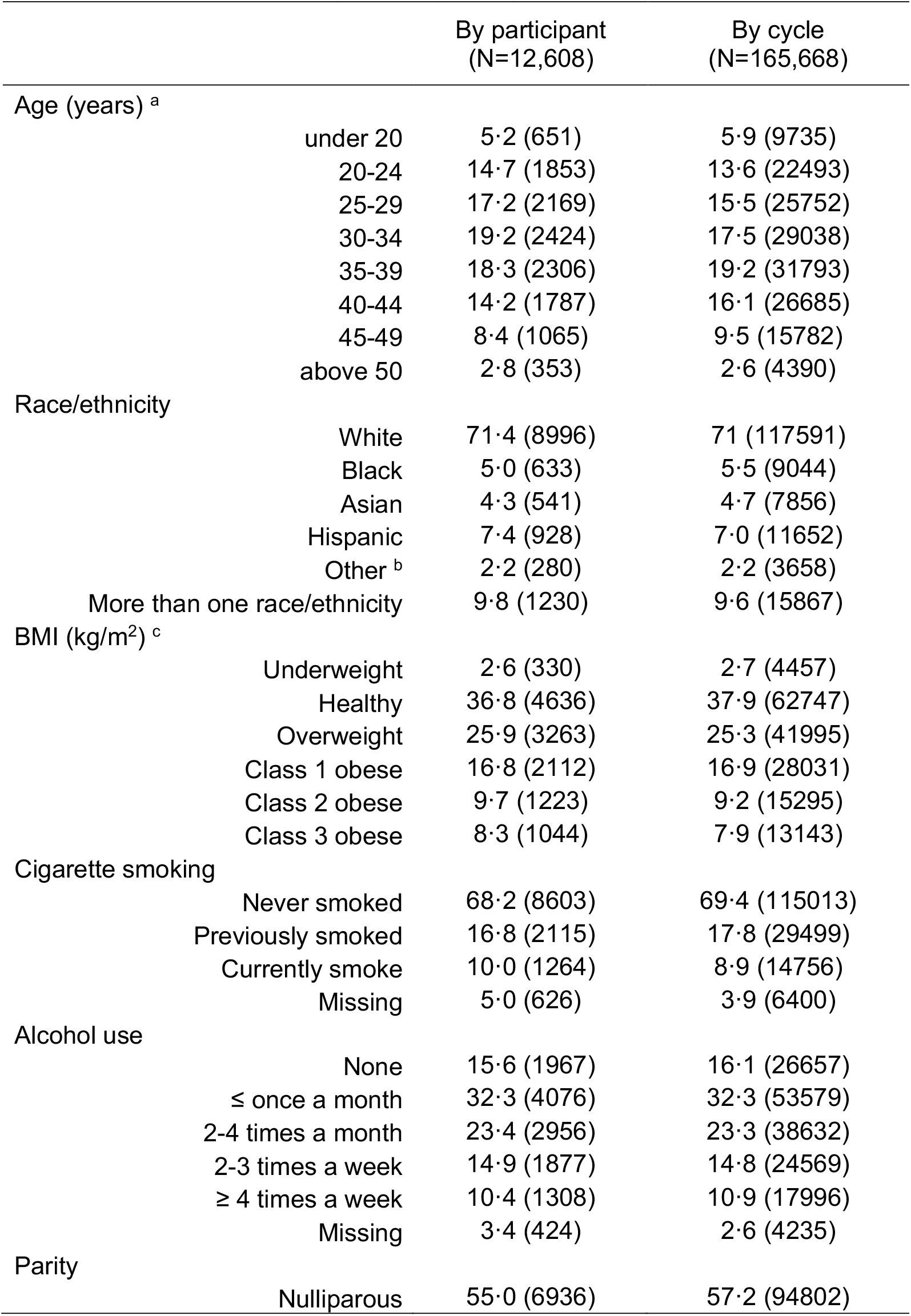
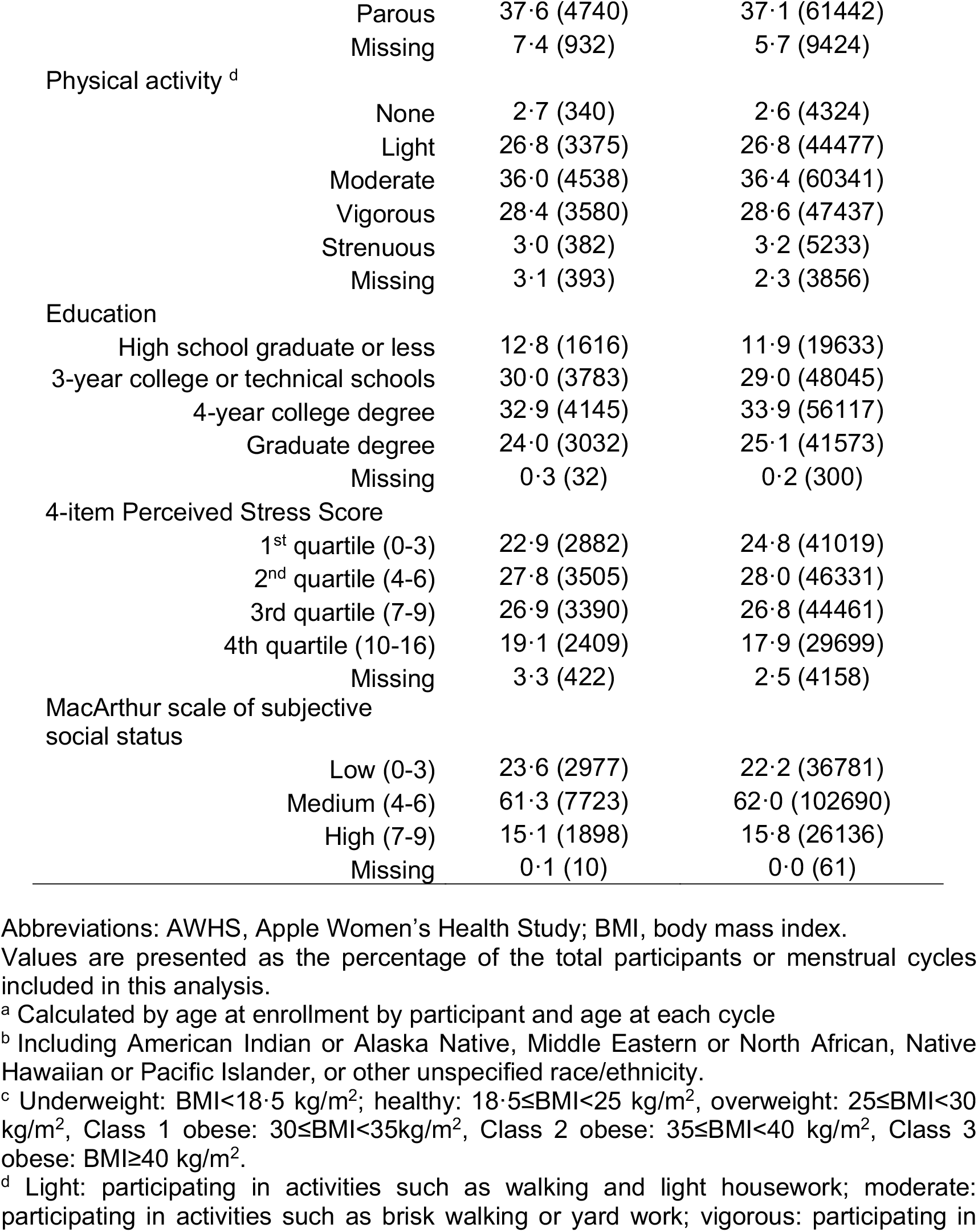
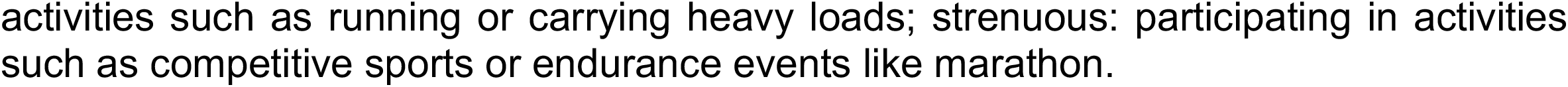
Distribution (% and N) of key characteristics in AWHS.

After adjusting for all covariates, we found mean menstrual cycle length to differ by age, race/ethnicity, and BMI groups (Table 2). Results for age were presented using age 35-39 as the reference because this group had the lowest cycle variability. Compared to the referent group, the mean cycle length was 1·6 (95%CI: 1·3, 1·9), 1·4 (1·2, 1·7), 1·1 (0·9, 1·3) and 0·6 (0·4, 0·7) days longer for women aged under 20, 20-24, 25-29, and 30-34, respectively. Menstrual cycle length continued to decrease by 0·5 (−0·3, 0·7) and 0·3 (−0·1, 0·6) days in the 40-44 and 45-49 age groups, respectively, and increased by 2·0 (1·6, 2·4) days among participants above age 50. Compared to White participants, the cycles of Asian participants were 1·6 (1·2, 2·0) days longer and the cycles for Hispanic participants were 0·7 (0·4, 1·0) days longer. No notable differences were found among the remaining race/ethnicity groups, with cycle lengths were 0·2 (−0·1, 0·6) days shorter in Black participants, and were 0·2 (−0·4, 0·7) and 0·1 (−0·2, 0·4) days longer for those in the other race/ethnicity group and who reported more than one race/ethnicity. Compared to those with healthy BMI, the cycles of those with overweight were 0·3 (0·1, 0·4) days longer, with Class 1 obesity were 0·5 (0·3, 0·8) days longer, with Class 2 obesity were 0·8 (0·5, 1·0) days longer, and Class 3 obesity were 1·5 (1·2, 1·8) days longer. The linear quantile mixed model suggested similar differences of cycle length by age, race/ethnicity, and BMI except that some patterns were more evident for cycle length on the 75^th^ percentile than the median and on the 25^th^ percentile (Table 2, Figure 1), reflecting the skewed distribution of cycle length. No notable effect modification was found between age and race/ethnicity and between age and BMI (p for interaction = 0·31 and 0·25). Obesity was associated with longer mean cycle length in White and Hispanic participants, while the associations were more evident for the Hispanic group (p for interaction = 0·0033). An increase in cycle length with obesity for Black participants was not apparent in the mean or median estimates but was evident when the 75^th^ percentile was examined. The differences of cycle length by BMI were moderate and statistically null in Asian participants (Figure 2).

**Table 2.**
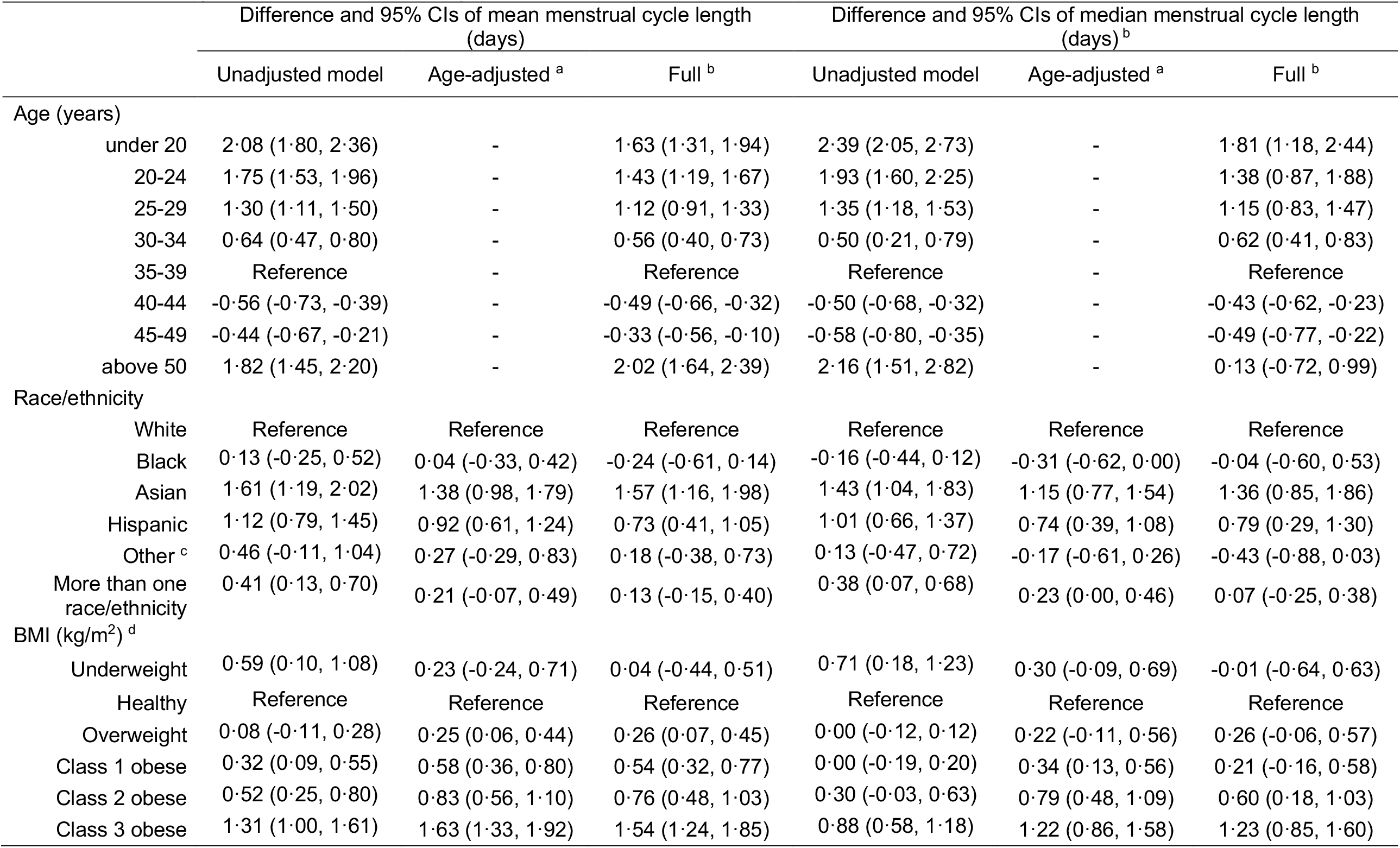
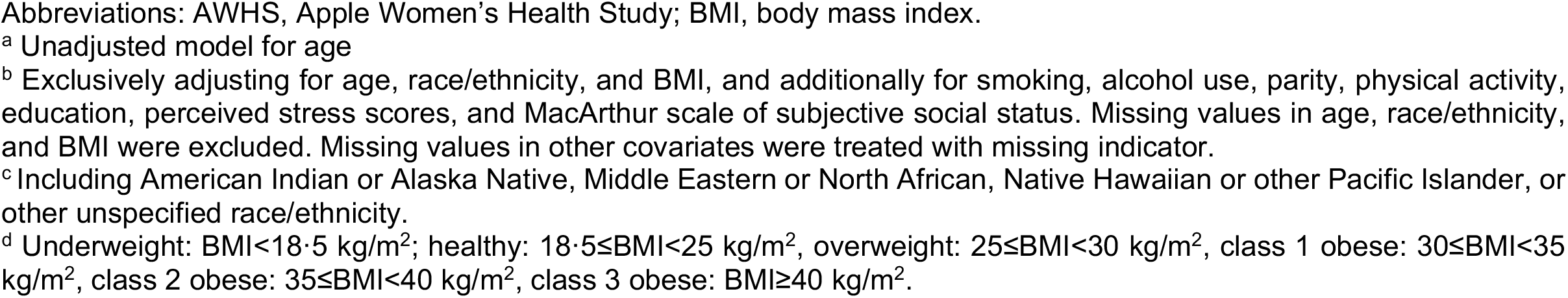
Differences and 95% confidence intervals (95%CIs) of mean menstrual cycle length with age, race/ethnicity, and BMI in 165,668 cycles from 12,608 participants in the AWHS.

|  | Difference and 95% CIs of mean menstrual cycle length<br>(days) |  |  | Difference and 95% CIs of median menstrual cycle length<br>(days)<sup>b</sup> |  |  |
| --- | --- | --- | --- | --- | --- | --- |
|  | Unadjusted model | Age-adjusted<sup>a</sup> | Full<sup>b</sup> | Unadjusted model | Age-adjusted<sup>a</sup> | Full<sup>b</sup> |
| **Age (years)** |  |  |  |  |  |  |
| under 20 | 2.08 (1.80, 2.36) | - | 1.63 (1.31, 1.94) | 2.39 (2.05, 2.73) | - | 1.81 (1.18, 2.44) |
| 20-24 | 1.75 (1.53, 1.96) | - | 1.43 (1.19, 1.67) | 1.93 (1.60, 2.25) | - | 1.38 (0.87, 1.88) |
| 25-29 | 1.30 (1.11, 1.50) | - | 1.12 (0.91, 1.33) | 1.35 (1.18, 1.53) | - | 1.15 (0.83, 1.47) |
| 30-34 | 0.64 (0.47, 0.80) | - | 0.56 (0.40, 0.73) | 0.50 (0.21, 0.79) | - | 0.62 (0.41, 0.83) |
| 35-39 | Reference | - | Reference | Reference | - | Reference |
| 40-44 | -0.56 (-0.73, -0.39) | - | -0.49 (-0.66, -0.32) | -0.50 (-0.68, -0.32) | - | -0.43 (-0.62, -0.23) |
| 45-49 | -0.44 (-0.67, -0.21) | - | -0.33 (-0.56, -0.10) | -0.58 (-0.80, -0.35) | - | -0.49 (-0.77, -0.22) |
| above 50 | 1.82 (1.45, 2.20) | - | 2.02 (1.64, 2.39) | 2.16 (1.51, 2.82) | - | 0.13 (-0.72, 0.99) |
| **Race/ethnicity** |  |  |  |  |  |  |
| White | Reference | Reference | Reference | Reference | Reference | Reference |
| Black | 0.13 (-0.25, 0.52) | 0.04 (-0.33, 0.42) | -0.24 (-0.61, 0.14) | -0.16 (-0.44, 0.12) | -0.31 (-0.62, 0.00) | -0.04 (-0.60, 0.53) |
| Asian | 1.61 (1.19, 2.02) | 1.38 (0.98, 1.79) | 1.57 (1.16, 1.98) | 1.43 (1.04, 1.83) | 1.15 (0.77, 1.54) | 1.36 (0.85, 1.86) |
| Hispanic | 1.12 (0.79, 1.45) | 0.92 (0.61, 1.24) | 0.73 (0.41, 1.05) | 1.01 (0.66, 1.37) | 0.74 (0.39, 1.08) | 0.79 (0.29, 1.30) |
| Other<sup>c</sup> | 0.46 (-0.11, 1.04) | 0.27 (-0.29, 0.83) | 0.18 (-0.38, 0.73) | 0.13 (-0.47, 0.72) | -0.17 (-0.61, 0.26) | -0.43 (-0.88, 0.03) |
| More than one race/ethnicity | 0.41 (0.13, 0.70) | 0.21 (-0.07, 0.49) | 0.13 (-0.15, 0.40) | 0.38 (0.07, 0.68) | 0.23 (0.00, 0.46) | 0.07 (-0.25, 0.38) |
| **BMI (kg/m<sup>2</sup>)<sup>d</sup>** |  |  |  |  |  |  |
| Underweight | 0.59 (0.10, 1.08) | 0.23 (-0.24, 0.71) | 0.04 (-0.44, 0.51) | 0.71 (0.18, 1.23) | 0.30 (-0.09, 0.69) | -0.01 (-0.64, 0.63) |
| Healthy | Reference | Reference | Reference | Reference | Reference | Reference |
| Overweight | 0.08 (-0.11, 0.28) | 0.25 (0.06, 0.44) | 0.26 (0.07, 0.45) | 0.00 (-0.12, 0.12) | 0.22 (-0.11, 0.56) | 0.26 (-0.06, 0.57) |
| Class 1 obese | 0.32 (0.09, 0.55) | 0.58 (0.36, 0.80) | 0.54 (0.32, 0.77) | 0.00 (-0.19, 0.20) | 0.34 (0.13, 0.56) | 0.21 (-0.16, 0.58) |
| Class 2 obese | 0.52 (0.25, 0.80) | 0.83 (0.56, 1.10) | 0.76 (0.48, 1.03) | 0.30 (-0.03, 0.63) | 0.79 (0.48, 1.09) | 0.60 (0.18, 1.03) |
| Class 3 obese | 1.31 (1.00, 1.61) | 1.63 (1.33, 1.92) | 1.54 (1.24, 1.85) | 0.88 (0.58, 1.18) | 1.22 (0.86, 1.58) | 1.23 (0.85, 1.60) |

Abbreviations: AWHHS, Apple Women's Health Study; BMI, body mass index.
<sup>a</sup> Unadjusted model for age
<sup>b</sup> Exclusively adjusting for age, race/ethnicity, and BMI, and additionally for smoking, alcohol use, parity, physical activity, education, perceived stress scores, and MacArthur scale of subjective social status. Missing values in age, race/ethnicity, and BMI were excluded. Missing values in other covariates were treated with missing indicator.
<sup>c</sup> Including American Indian or Alaska Native, Middle Eastern or North African, Native Hawaiian or other Pacific Islander, or other unspecified race/ethnicity.
<sup>d</sup> Underweight: $\text{BMI} < 18.5 \text{ kg/m}^2$ ; healthy: $18.5 \leq \text{BMI} < 25 \text{ kg/m}^2$ , overweight: $25 \leq \text{BMI} < 30 \text{ kg/m}^2$ , class 1 obese: $30 \leq \text{BMI} < 35 \text{ kg/m}^2$ , class 2 obese: $35 \leq \text{BMI} < 40 \text{ kg/m}^2$ , class 3 obese: $\text{BMI} \geq 40 \text{ kg/m}^2$ .

**Figure 1.**
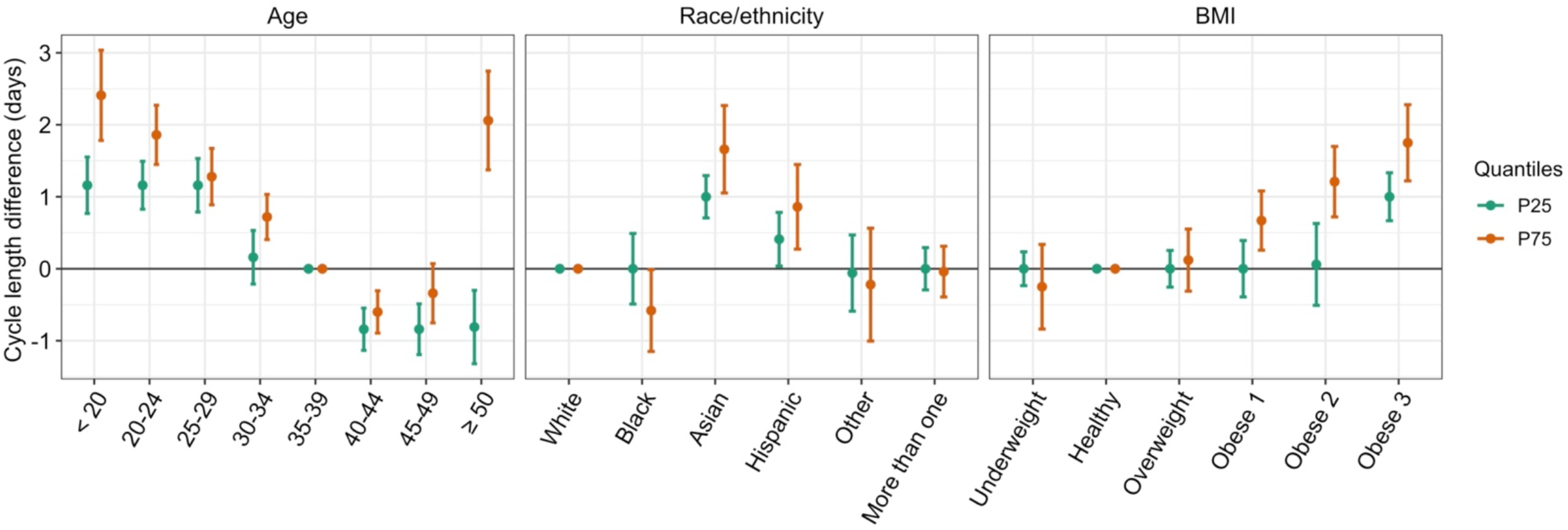
Differences and 95%CIs of menstrual cycle length on the 25^th^ and 75^th^ percentiles by age, race/ethnicity, and BMI. Abbreviation: CIs, confidence intervals; BMI, body mass index, P25: 25^th^ percentile; P75: 75^th^ percentile. Exclusively adjusting for age, race/ethnicity, and BMI, and additionally for smoking, alcohol drinking, parity, physical activity, education, perceived stress scores, and MacArthur scale of subjective social status. Missing values in age, race/ethnicity, and BMI were excluded. Missing values in other covariates were treated with missing indicator. The other race/ethnicity group includes American Indian or Alaska Native, Middle Eastern or North African, Native Hawaiian or other Pacific Islander, or other unspecified race/ethnicity. Underweight: BMI<18·5 kg/m^2^; healthy: 18·5≤BMI<25 kg/m^2^, overweight: 25≤BMI<30 kg/m^2^, class 1 obese: 30≤BMI<35 kg/m^2^, class 2 obese: 35≤BMI<40 kg/m^2^, class 3 obese: BMI≥40 kg/m^2^.

**Figure 2.**
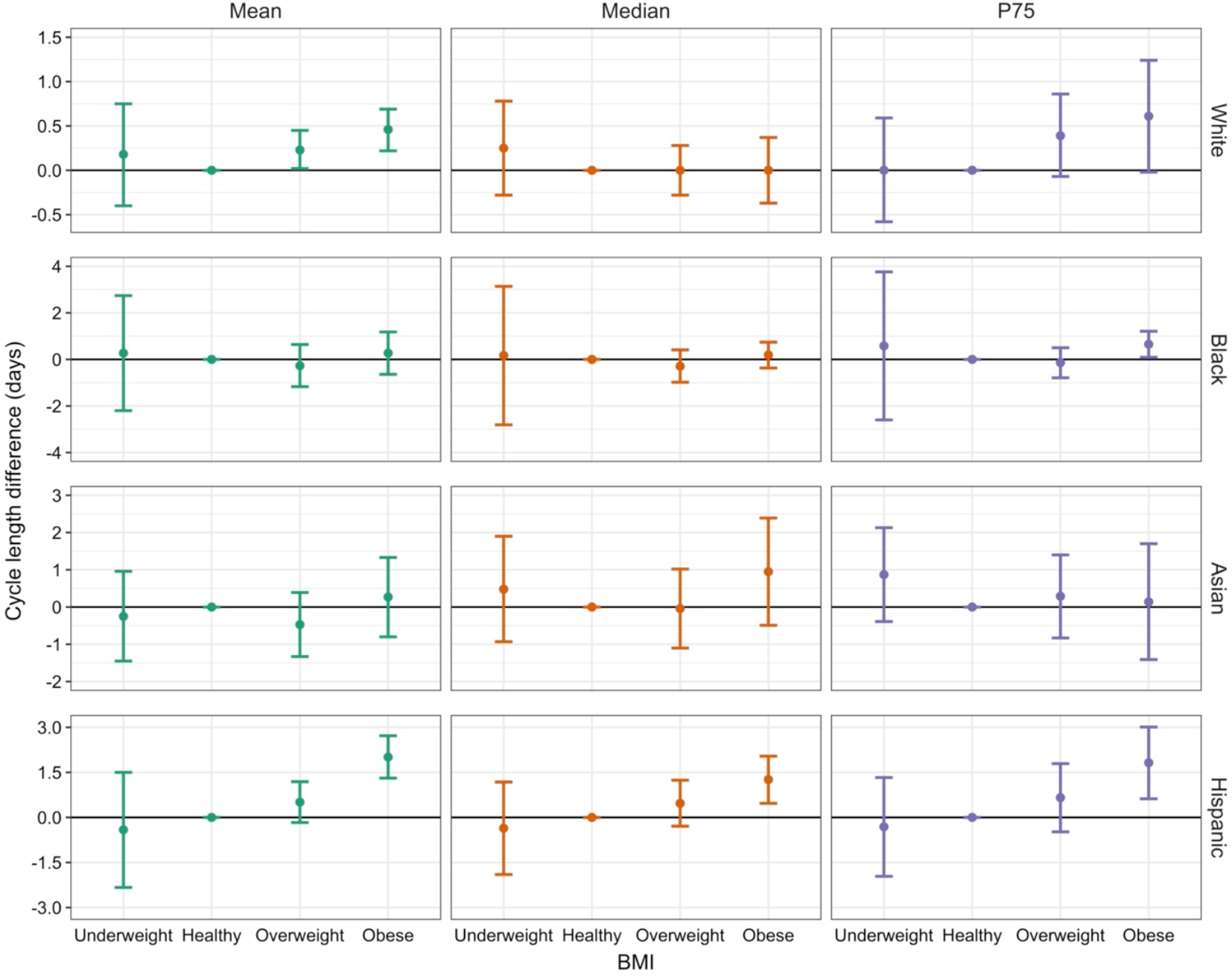
Differences and 95%CIs of the mean and median menstrual cycle length with BMI by race/ethnicity groups. Abbreviation: CIs, confidence intervals; BMI, body mass index Adjusting for age, smoking, alcohol drinking, parity, physical activity, education, perceived stress scores, and MacArthur scale of subjective social status. Missing values in age, race/ethnicity, and BMI were excluded. Missing values in other covariates were treated with missing indicator. Analysis was restricted to participants who were under age 50 years and had BMI<40 kg/m^2^, and to White, Black, Asian, and Hispanic participants to avoid having strata with few observations. Underweight: BMI<18·5 kg/m^2^; healthy: 18·5≤BMI<25 kg/m^2^, overweight: 25≤BMI<30 kg/m^2^, obese: 30≤BMI<40 kg/m^2^.

The odds of long or short cycles also differed by age, race/ethnicity, and BMI (Table 3). Younger participants were more likely than those aged 35-39 years to experience long cycles and less likely to have short cycles, while those in 45-49 year age group were more likely to experience both long and short cycles. This trend was more evident in participants above age 50 years. Asian and Hispanic participants were more likely to have long cycles and less likely to have short cycles compared to White participants. Similarly, obese participants were more likely to have a long cycle but not a short cycle (Table 3). All sensitivity analyses showed similar results to the main analyses (Table S3-S7).

**Table 3.**
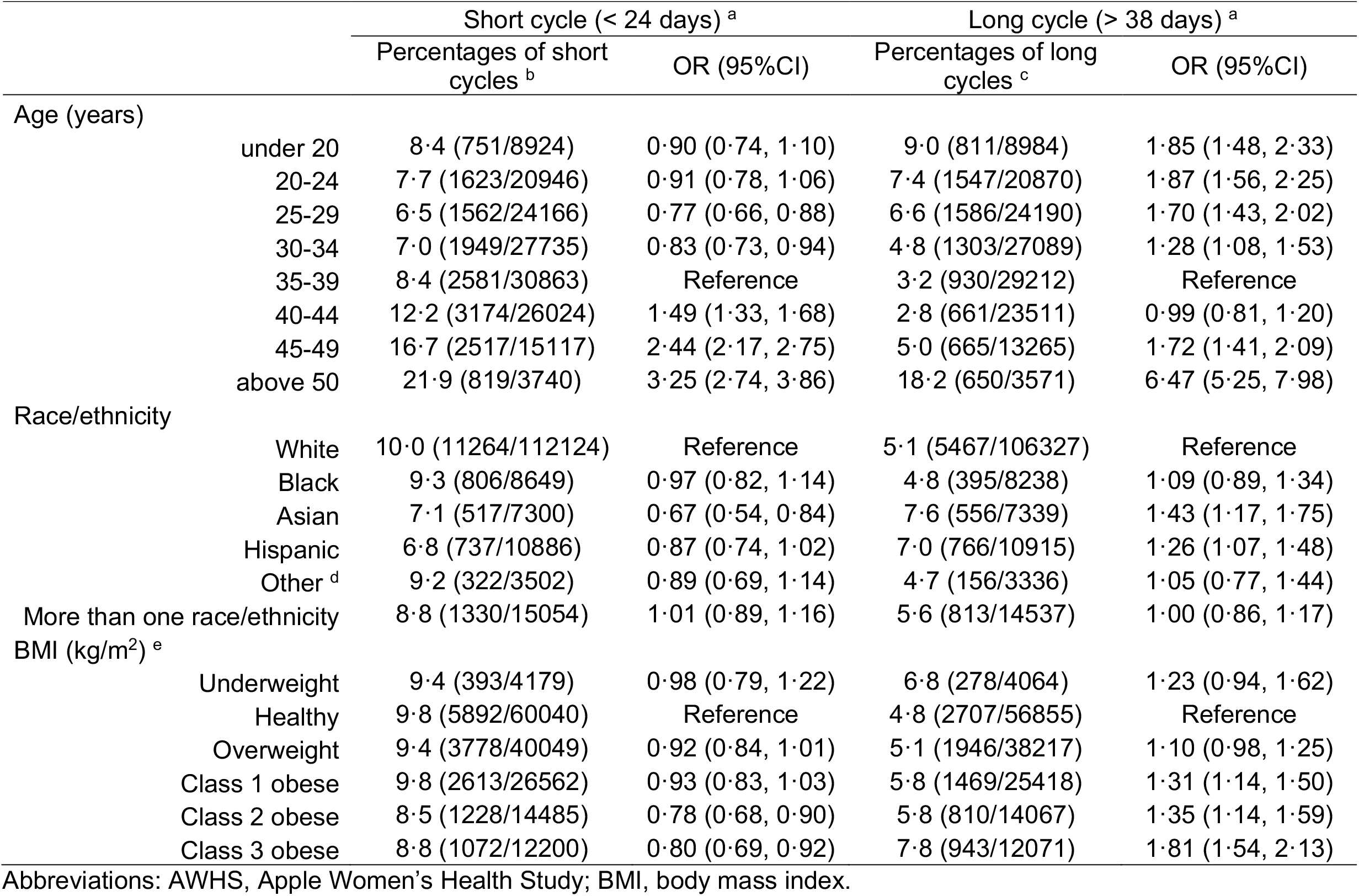
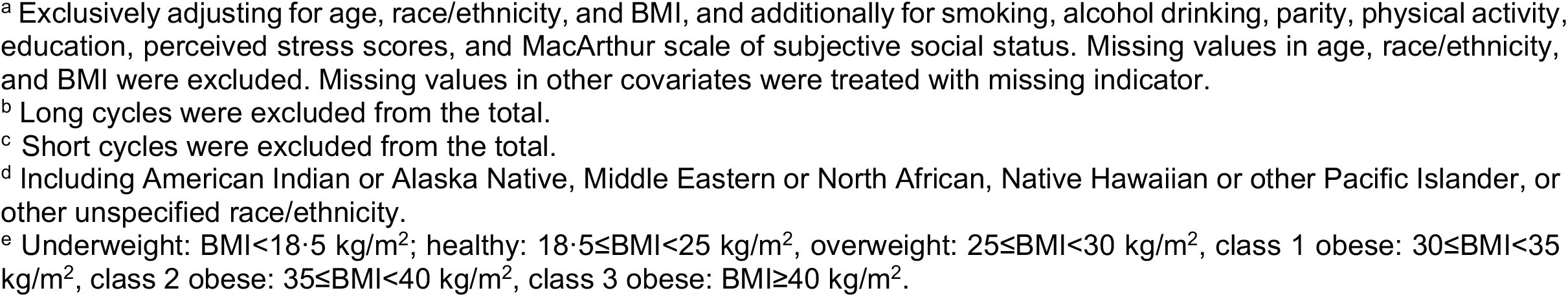
Odds ratios (ORs) and 95% confidence intervals (95%CIs) of experiencing a short (< 24 days) or long (> 38 days) menstrual cycle by age, race/ethnicity, and BMI in 165,668 cycles from 12,608 participants in the AWHS.

|  |  | Short cycle (< 24 days) <sup>a</sup> |  | Long cycle (> 38 days) <sup>a</sup> |  |
| --- | --- | --- | --- | --- | --- |
|  |  | Percentages of short cycles <sup>b</sup> | OR (95%CI) | Percentages of long cycles <sup>c</sup> | OR (95%CI) |
| Age (years) |  |  |  |  |  |
|  | under 20 | 8.4 (751/8924) | 0.90 (0.74, 1.10) | 9.0 (811/8984) | 1.85 (1.48, 2.33) |
|  | 20-24 | 7.7 (1623/20946) | 0.91 (0.78, 1.06) | 7.4 (1547/20870) | 1.87 (1.56, 2.25) |
|  | 25-29 | 6.5 (1562/24166) | 0.77 (0.66, 0.88) | 6.6 (1586/24190) | 1.70 (1.43, 2.02) |
|  | 30-34 | 7.0 (1949/27735) | 0.83 (0.73, 0.94) | 4.8 (1303/27089) | 1.28 (1.08, 1.53) |
|  | 35-39 | 8.4 (2581/30863) | Reference | 3.2 (930/29212) | Reference |
|  | 40-44 | 12.2 (3174/26024) | 1.49 (1.33, 1.68) | 2.8 (661/23511) | 0.99 (0.81, 1.20) |
|  | 45-49 | 16.7 (2517/15117) | 2.44 (2.17, 2.75) | 5.0 (665/13265) | 1.72 (1.41, 2.09) |
|  | above 50 | 21.9 (819/3740) | 3.25 (2.74, 3.86) | 18.2 (650/3571) | 6.47 (5.25, 7.98) |
| Race/ethnicity |  |  |  |  |  |
|  | White | 10.0 (11264/112124) | Reference | 5.1 (5467/106327) | Reference |
|  | Black | 9.3 (806/8649) | 0.97 (0.82, 1.14) | 4.8 (395/8238) | 1.09 (0.89, 1.34) |
|  | Asian | 7.1 (517/7300) | 0.67 (0.54, 0.84) | 7.6 (556/7339) | 1.43 (1.17, 1.75) |
|  | Hispanic | 6.8 (737/10886) | 0.87 (0.74, 1.02) | 7.0 (766/10915) | 1.26 (1.07, 1.48) |
|  | Other <sup>d</sup> | 9.2 (322/3502) | 0.89 (0.69, 1.14) | 4.7 (156/3336) | 1.05 (0.77, 1.44) |
|  | More than one race/ethnicity | 8.8 (1330/15054) | 1.01 (0.89, 1.16) | 5.6 (813/14537) | 1.00 (0.86, 1.17) |
| BMI (kg/m<sup>2</sup>) <sup>e</sup> |  |  |  |  |  |
|  | Underweight | 9.4 (393/4179) | 0.98 (0.79, 1.22) | 6.8 (278/4064) | 1.23 (0.94, 1.62) |
|  | Healthy | 9.8 (5892/60040) | Reference | 4.8 (2707/56855) | Reference |
|  | Overweight | 9.4 (3778/40049) | 0.92 (0.84, 1.01) | 5.1 (1946/38217) | 1.10 (0.98, 1.25) |
|  | Class 1 obese | 9.8 (2613/26562) | 0.93 (0.83, 1.03) | 5.8 (1469/25418) | 1.31 (1.14, 1.50) |
|  | Class 2 obese | 8.5 (1228/14485) | 0.78 (0.68, 0.90) | 5.8 (810/14067) | 1.35 (1.14, 1.59) |
|  | Class 3 obese | 8.8 (1072/12200) | 0.80 (0.69, 0.92) | 7.8 (943/12071) | 1.81 (1.54, 2.13) |

Abbreviations: AWHs, Apple Women's Health Study; BMI, body mass index.
<sup>a</sup> Exclusively adjusting for age, race/ethnicity, and BMI, and additionally for smoking, alcohol drinking, parity, physical activity, education, perceived stress scores, and MacArthur scale of subjective social status. Missing values in age, race/ethnicity, and BMI were excluded. Missing values in other covariates were treated with missing indicator.
<sup>b</sup> Long cycles were excluded from the total.
<sup>c</sup> Short cycles were excluded from the total.
<sup>d</sup> Including American Indian or Alaska Native, Middle Eastern or North African, Native Hawaiian or other Pacific Islander, or other unspecified race/ethnicity.
<sup>e</sup> Underweight: $\text{BMI} < 18.5 \text{ kg/m}^2$ ; healthy: $18.5 \leq \text{BMI} < 25 \text{ kg/m}^2$ , overweight: $25 \leq \text{BMI} < 30 \text{ kg/m}^2$ , class 1 obese: $30 \leq \text{BMI} < 35 \text{ kg/m}^2$ , class 2 obese: $35 \leq \text{BMI} < 40 \text{ kg/m}^2$ , class 3 obese: $\text{BMI} \geq 40 \text{ kg/m}^2$ .

The average within-individual variability varied between 4-6 days across most of the age, race/ethnicity, and BMI groups. Changes in cycle variability with age were the most evident compared to race/ethnicity and BMI (Table 4). The 35-39 year age group had the lowest variability and relative increases in variability were observed in both younger and older age groups. For example, compared to the 35-39 age group, participants in the under 20, 20-24, 25-29, and 30-34 age groups had 45% (43, 48), 37% (34, 40), 25% (22, 28), and 13% (10, 16) higher cycle variability, respectively, and those who aged between 40-44 had 45-49 had 6% (1, 10) and 44% (40, 49) higher variability. In addition, participants over age 50 years had an estimated 200% (191, 209) higher variability compared to those age 35-39 years. Asian and Hispanic participants had 9% (3, 15) and 10% (3, 17) higher cycle variability compared to White participants, while no notable differences in cycle variability were found for those who were Black (−3%, 95%CI= -5, 0), who reported other race/ethnicity (−4%, 95%CI= -9, 0), and who reported more than one race/ethnicity (−5%, 95%CI=-15, 4) compared to White participants. Obese participants had larger within-individual variability than those with healthy BMI, with increases of 12% (10, 15), 10% (6, 13), and 27% (23, 30) for those with Class 1, 2, and 3 obesity. The ORs for irregularity showed similar patterns of associations by age, race/ethnicity, and BMI groups as those for cycle variability (Table 4). Results for irregularity were similar when applying different definitions (Table S8).

**Table 4.**
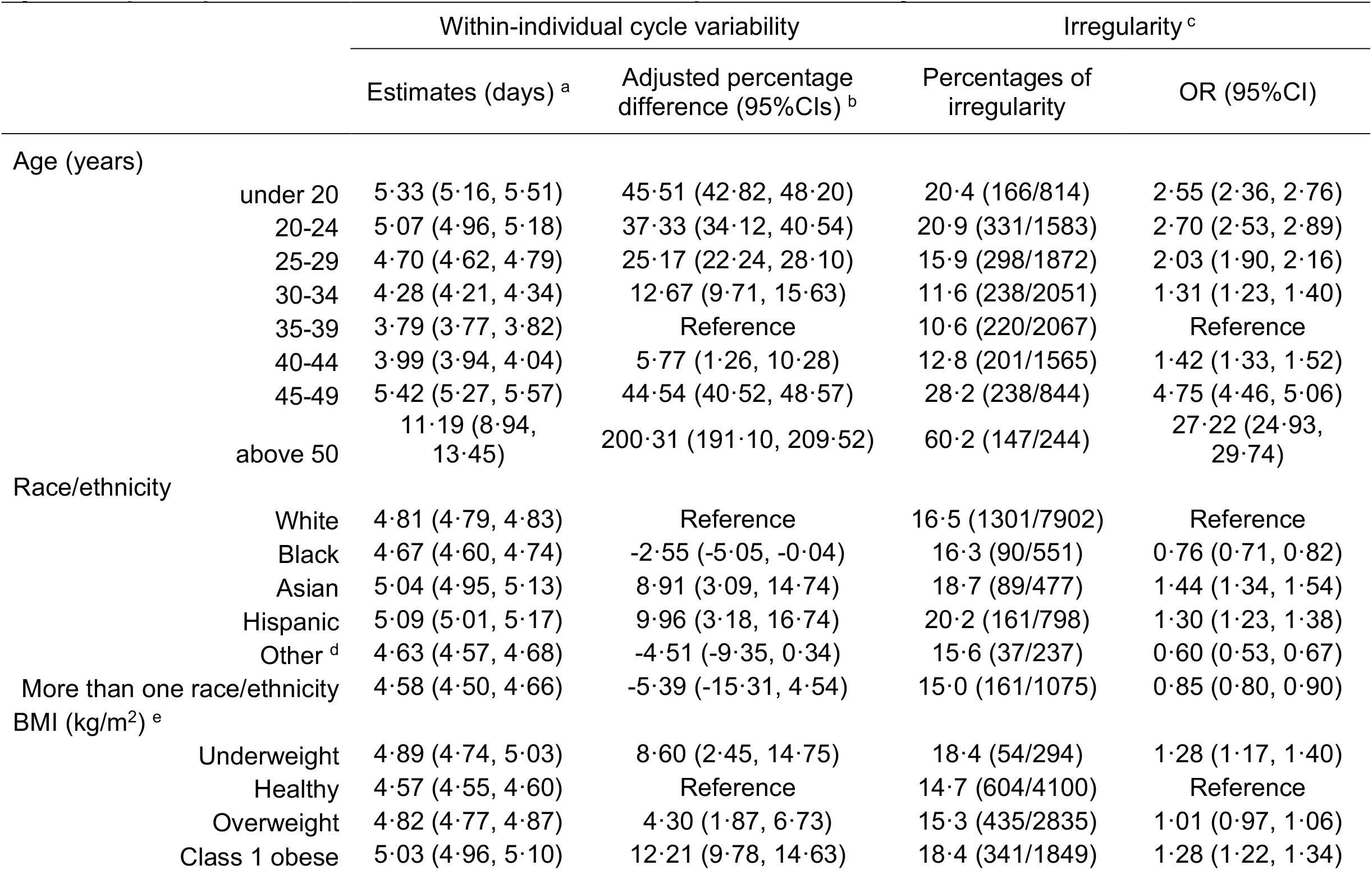
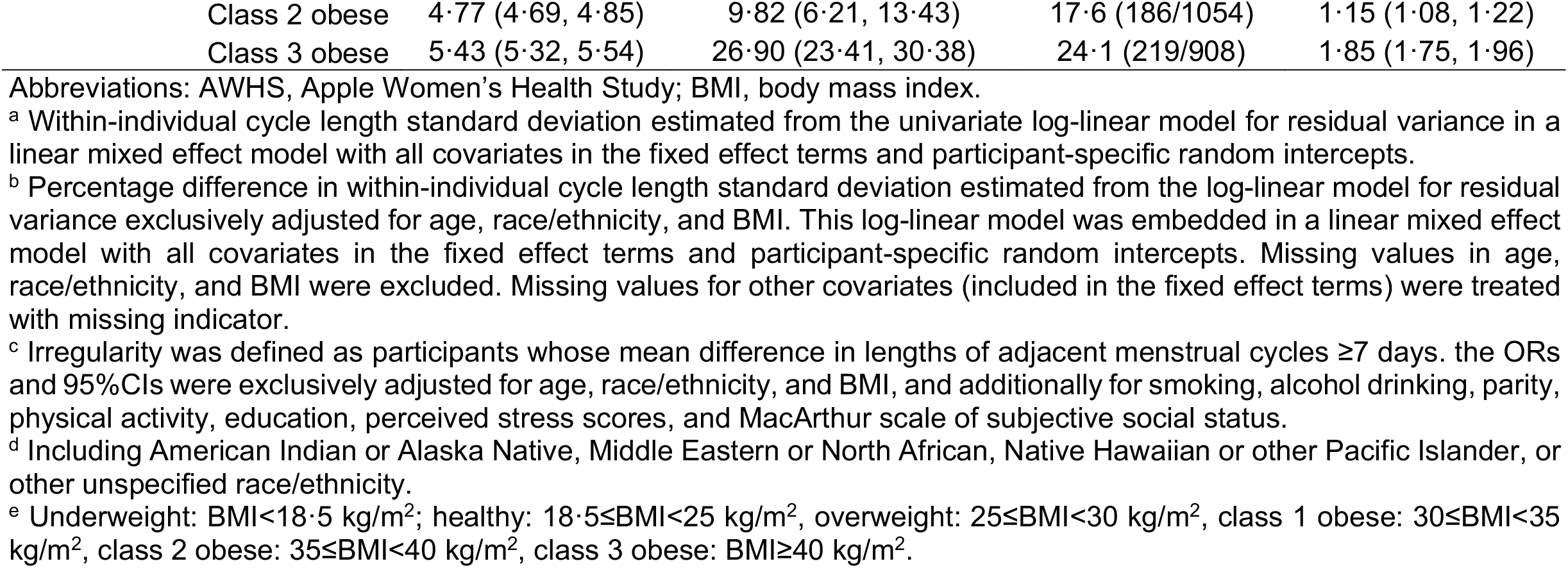
Estimates and percentage change of within-individual standard deviations (SDs), and odds ratios (ORs) of experiencing irregularity with 95% confidence intervals (95%CIs) by age, race/ethnicity, and BMI in 163,275 cycles by 11,040 participants who contributed at least three complete menstrual cycles in the AWHS.

## Discussion

In this digital cohort study, we comprehensively examined differences in menstrual cycle length and variability by age, race/ethnicity, and BMI. Our results for age suggest that compared to participants in early reproductive years (i.e., under age 20 or between age 20-24), menstrual cycles were shorter for those in the older age groups up until age 50 years. Cycle length variability was also smaller in older reproductive age groups but became considerably larger among those at age 45-49 years and 50 years and above. Asian and Hispanic participants and those with higher BMI had longer cycles and higher cycle variability.

Compared to the 5-95^th^ percentiles of cycle length given by FIGO as the normal range (24-38 days), cycle length distribution in AWHS showed a comparable 95^th^ percentile but a shorter 5^th^ percentile (23). Our findings on differences in cycle length and variability by age were consistent with previous reports using data from diaries (7,8,12,40) and mobile apps (21,22,39), and aligned with the menstrual cycle characteristics across reproductive life span (41). The observed shorter cycle length in older groups under age 50 may be explained by the decreasing ovarian reserve over time, as previous studies have showed lower ovarian reserve was associated with shorter cycle length in reproductive aged women (42,43). We found changes in the mean and median cycle length differed for those aged above 50 years. Participants who were over age 50 years had much longer and more variable cycles, as would be expected during the late menopausal transition that is characterized by highly variable cycles and frequent anovulation (41), although studies of menstrual cycle characteristics in late menopausal stage are still limited.

Across all race/ethnicity groups, Asian and Hispanic participants had longer cycle lengths, moderately larger within-individual cycle variability, and were more likely to experience cycle irregularity. No notable differences in cycle length and variability were found between Black and White participants. Studies directly comparing of menstrual patterns across race and ethnicity groups were scarce. One study in African- and European-American adolescents reported African-American girls had moderately shorter cycle length (11). Two studies among peri-menopausal women in the US found Asians (Japanese and Chinese) had longer cycles compared to the White participants and Hispanic individuals were more likely to experience a cycle longer than 33 days, while no notable differences were observed between African-Americans and those who were White (16,32). A study of reproductive aged women in the semiconductor industry also showed longer cycle length in Asians compared to those who were White (14). A recent app-based study reported longer and more irregular cycles for US Hispanic users compared to those who were Black (39). Previous studies have linked higher follicular phase estrogen with shorter menstrual cycle length (44–46). Other studies, however, reported higher estrogen levels in African-Americans, Hispanics, and Asians compared to those who were White, but the timing of hormone measurement within the menstrual cycle was different across these studies (28–30). Ovarian reserve can be an underlying factor for the observed differences. It has been documented in the literature that Hispanic and Asian women had higher anti-Müllerian hormone, a marker of ovarian reserve, compared to White females, but evidence was mixed for African-Americans (31). Therefore, differences in ovarian reserve may not fully explain the observed differences of menstrual cycle length by race/ethnicity. There are several factors that can interfere with the hypothalamus-pituitary-ovary axis, thyroids, adrenal gland, and, consequentially, menstrual cycles, such as BMI, physical activity, stress, and socioeconomic status. However, the racial and ethnic differences of cycle length persisted after we controlled for these factors in the model. Therefore, our results may be explained by other unmeasured factors such as disparities in life-course exposures to cultural, social, and environmental factors of menstrual and reproductive health, which warrants further investigation (12,17).

Our results also suggest overweight and obese participants have longer menstrual cycles, greater cycle variability, and are more likely to experience irregularity than the healthy weight participants. Previous studies have linked higher body weight with long menstrual cycles and higher cycle variability (10,12,13). A study among a very large number of app users reported participants with BMI between 35-50 kg/m^2^ had higher cycle variation by 0·4 days and longer mean cycle length by 0·5 days than those with a healthy BMI (21). However, only 8% of the participants had BMI above 30 kg/m^2^. Another large app-based study found small differences in cycle length and variability across BMI groups overall, but it was the underweight group that stood out: they had higher cycle variability and a higher proportion of participants with BMI≥35 kg/m^2^ had a median cycle length above 36 days (22). Obesity has been linked with endocrine disruptions such as hyperinsulinemia and excess leptin secretion, which may affect the hormonal regulation of menstrual function (47). Other pathways include obesity-related chronic low-grade inflammation and oxidative stress, which can adversely affect the ovary. Fat tissue is a peripheral producer of estrogen (estrone), which can affect the regulatory activity of the hypothalamic-pituitary-ovarian axis and therefore, possibly inhibits ovarian gonadotropin and estrogen production (48,49). Obesity and long and/or irregular menstrual cycles are individually and jointly associated with cardiometabolic risk (3,4), which has been found disproportionally higher among Hispanic and Black females in the US (50). However, the associations of obesity with longer cycles were stronger in Hispanic but not in Black participants, though both had relatively small sample size in our study. In addition, we excluded participants with Class 3 obesity group in the effect modification analysis to avoid subgroups with small sample sizes, which limited our ability to fully quantify the heterogenous associations of BMI and cycle length by race/ethnicity. Future studies could explore the possible interactions among race/ethnicity, obesity, and menstrual function to better understand their relationships with cardiometabolic health.

Though we had sufficient statistical power to detect minor differences in menstrual cycle length, our sample size was relatively small compared to other studies using menstrual tracking app data (18,19,21,22,39). A notable limitation of this study is the reliance on self-reported/tracking information to identify menstrual cycles and measure the covariates, with the user’s reporting/tracking behavior and health conditions possibly affecting accuracy of the study data. There is no widely accepted gold standard in menstrual cycle measurement in epidemiological studies. Cycle data collected using prospective diary is considered more accurate than the self-reported typical cycle length and variability in surveys (51–53). However, inaccurate reporting is still possible in a prospective diary. Previous studies have suggested that when height and weight are self-reported, use of the continuous measure of BMI calculated from these values could introduce less bias to the model estimates compared to the categorization of BMI (54,55). Our sensitivity analysis using continuous BMI measures showed estimates similar to those obtained from our primary analyses (Table S9). In addition, although BMI-defined obesity is a common and convenient metric to identify excess body fat that can adversely affect health, it could lead to misclassification, and the degree and direction will likely differ by race/ethnicity and other factors such as age and education (56,57). We did not have data on reproductive hormones or ovulation testing, which limited our ability to consider more detailed aspects of menstruation. When comparing cycle characteristics across age groups, we arbitrarily used the 35-39 year age group as the reference because this group had the lowest cycle variability. Therefore, the estimates should be interpreted with considerations on the reproductive life stages to which these age groups correspond. Part of our study data were collected during the COVID-19 pandemic. Our sensitivity analysis among participants who never had known COVID-19 infection showed comparable estimates to those in the main model. In addition, an on-going analysis in AWHS suggests moderate and very short-term changes in menstrual cycle length associated with COVID-19 vaccination (58)(Preprint). Therefore, we believed both COVID-19 infection and vaccination had minimal impact on our results. Generalizability is also limited because our study participants are all iPhone users and those who can communicate in English, which could lead to underrepresentation of individuals with low socioeconomic status and Hispanic population.

This study examined menstrual cycle length and variability by age, race/ethnicity, and BMI using data collected from mobile apps in a large, diverse population in the US. Our findings confirmed previous observations of associations between age and BMI and cycle length and variability. Asian and Hispanic participants had longer menstrual cycle length and higher variability than the other groups. Future studies are needed to confirm the observed racial and ethnic differences, explore whether these menstrual differences account for reproductive health disparities, and identify the underlying determinants that result in menstrual cycle differences.

## Data Availability

Aggregated deidentified data that support the findings of this study may be available upon request from the corresponding author Dr. Shruthi Mahalingaiah. Any request for data will be evaluated and responded to in a manner consistent with policies intended to protect participant confidentiality and language in the Study protocol and informed consent form.

## Acknowledgement

This study received funding from Apple. Inc and was not funded by NIH. The funding source played no role in the analysis and interpretation of data, and in the decision to submit the paper for publication. Apple. Inc provided platforms and tools for data collection. We would like to thank all the AWHS participants for signing up for the study and contributing to the advancement of women’s health research. We would also like to acknowledge Alexis de Figueiredo Veiga, Nicola Gallagher (former study team member at the Harvard T.H. Chan School of Public Health), Ariel Scalise, Carol Mita, and Gowthan Asokan for their work in supporting the study. AMZJ, DDB, and AJW are employed by NIH. Support for AMZJ, DDB, and AJW was provided by the Intramural Research Program of the National Institute of Environmental Health Sciences, National Institute of Health. BC, RH, JPO, and SM are in receipt of NIH grants.

## Contributors

HL was responsible for study design, data preparation, statistical analyses, interpretation of the results, manuscript writing, revision, and finalization. EAG contributed to interpretation of the results and manuscript revision. SM oversaw all aspects of the work and contributed to developing the concept, analysis plan, interpretation of the results, manuscript writing, and revision. BAC, RH, JPO, and MAW were responsible for study design, interpretation of the results, manuscript writing, and revision. AMZJ, DDB, and AJW provided critical reviews to the analysis and contributed to interpretation of the results and manuscript revision. CLC and TFC contributed to review of content in relation to the publication policy of Apple Inc. and did not participate in the analysis and interpretation of data. The corresponding author attests that all listed authors meet authorship criteria and that no others meeting the criteria have been omitted.

## Data sharing

Aggregated deidentified data that support the findings of this study may be available upon request from the corresponding author (SM). Any request for data will be evaluated and responded to in a manner consistent with policies intended to protect participant confidentiality and language in the Study protocol and informed consent form.

## Declaration of interests

All authors have completed the ICMJE uniform disclosure form at www.icmje.org/coi_disclosure.pdf and declare: no support from any organization for the submitted work; CLC and TFC are employed by Apple Inc. and own Apple Inc. stock. No financial relationships with any organizations that might have an interest in the submitted work in the previous three years; no other relationships or activities that could appear to have influenced the submitted work.

## Notes

### Competing Interest Statement

The authors have declared no competing interest.

### Author Declarations

This study has been approved by the Institutional Review Board at Advarra (CIRB #PRO00037562) and has been registered in Clinicaltrials.gov (NCT04196595).

