## Supplemental Materials for "Variation in menstrual cycle length by age, race/ethnicity, and body mass index in a large digital cohort of women in the US"

### Appendix

#### Table of contents


| | |
|---|---|
| Supplemental Methods | 1-6 |
| Table S1 Number of menstrual cycles by age and BMI across race/ethnicity groups in 165,668 menstrual cycles from 12,608 participants of AWHS | 7 |
| Table S2 Numbers of menstrual cycles across different cycle length groups by age, race/ethnicity, and BMI in 165,668 menstrual cycles from 12,608 participants of AWHS | 8 |
| Table S3 Differences and 95% confidence intervals (95%CIs) of mean menstrual cycle length with age, race/ethnicity, and BMI after restricting to participants who tracked ≥ 3 cycles, to participants under age 50, to cycles with confirmed accurate tracking, to cycles with complete data, and including participants with uterine fibroids | 9-10 |
| Table S4 Differences and 95% confidence intervals (95%CIs) of median menstrual cycle length with age, race/ethnicity, and BMI to participants who tracked ≥ 3 cycles, to participants under age 50, to cycles with confirmed accurate tracking, to cycles with complete data, and including participants with uterine fibroids | 11-12 |
| Table S5 Odds ratios (ORs) and 95% confidence intervals (95%CIs) of experiencing a short (<24 days) menstrual cycle by age, race/ethnicity, and BMI to participants who tracked ≥ 3 cycles, to participants under age 50, to cycles with confirmed accurate tracking, to cycles with complete data, and including participants with uterine fibroids | 13-14 |
| Table S6 Odds ratios (ORs) and 95% confidence intervals (95%CIs) of experiencing a long (>38 days) menstrual cycle by age, race/ethnicity, and BMI, after restricting to women who tracked ≥ 3 cycles, to women under age 50, to cycles with confirmed accurate tracking, to cycles with complete data, and including participants with uterine fibroids | 15-16 |
| Table S7 Differences and 95% confidence intervals (95%CIs) of mean menstrual cycle length with age, race/ethnicity, and BMI in 59,431 cycles from 4,119 AWHS participants who never had known COVID-19 infection | 17 |
| Table S8 Odds ratios (ORs) and 95% confidence intervals (95%CIs) of experiencing menstrual irregularity by age, race/ethnicity, and BMI using different criteria among 11,040 participants with ≥ 3 menstrual cycles in AWHS | 18-19 |
| Table S9 Differences and 95% confidence intervals of mean menstrual cycle length by BMI groups using categorical, continuous, and piecewise BMI | 20 |
| Figure S1 Inclusion and exclusion of menstrual cycles and AWHS participants | 21 |
| Figure S2 By-participant distribution (density and cumulative density) of menstrual cycle length across 165,668 cycles from 12,608 participants in the AWHS | 22 |

#### Supplemental Methods

##### Menstrual cycle identification

To minimize the influence of artifacts due to gaps in record-keeping, we excluded cycles that were atypically long and likely artifacts. These atypically long cycles were identified with an individual-specific threshold considering the typical menstrual cycle length and menstrual cycle variability of that individual. This approach was originally adopted from a large, population-based analysis using self-tracked cycle data from mobile apps and was modified based on our data [1]. Specifically, we used the median menstrual cycle length and median of the cycle length difference (calculated as the absolute value of the difference of the length between two adjacent cycles) to represent the typical menstrual cycle length and cycle variation of that person. We chose the median because the mean and standard deviation of cycle length in the raw data were more likely to be influenced by extreme values from artifacts. However, it is possible that a participant can naturally experience an atypically long cycle. Therefore, we added a 15-day window to the threshold to account for such possibility. We chose 15 days because our data suggested this as the optimal value to achieve a balance between identifying cycle artifacts and preserving natural cycle variation. The final individual-specific threshold was calculated as the sum of the median cycle length and median cycle length difference plus 15 days, and a cycle longer than this threshold value was identified as an artifact. Detecting cycle artifacts among peri-menopausal participants may be difficult because these individuals usually have large cycle variability and frequent anovulation [2,3]. Therefore, we only applied this identification in participants under age 50. For women above age 50, we included all their cycles into the analysis.

Among the 742,747 cycles within 10-90 days from 49,238 AWHS participants under age 50, a total of 29,174 cycles from 18,043 participants were identified as artifacts, which corresponds to an average of 1·62 cycles per individual. This average is comparable to the average of 1·59 cycles per user with artifacts in the previous study [1]. The distribution of median cycle length was largely unchanged before and after the exclusion, suggesting our approach only identified and excluded outliers for each indiviudal (Table SM1). As shown in Figure S1, there was a density peak for cycles that were approximately 21-32 days longer than median cycle length in the raw data, suggesting a possibility of cycle artifacts. After excluding these cycles, this atypical density peak disappeared. Additionally, the long right tail in the histogram after exclusion suggested the natural variability of menstrual cycle length was preserved. Figure SM2 compares the distribution of menstrual cycle length among cycles that were identified as artifacts and those that were not. Most cycles identified as artifacts were within 50-60 days long, which is approximately twice the length of a typical menstrual cycle. These atypically long cycles may be artificially created when a participant missed logging a bleeding period in this interval.

**Within cluster resampling for informative cluster size**

In analysis of clustered, correlated data, informative cluster size or non-ignorable cluster size can occur when the size of the cluster is informative of the outcome, which can lead to biased estimates when using marginal approaches such as the generalized estimating equations (GEEs) [4]. However, mixed effect models can avoid this bias [5]. Our preliminary analysis showed a negative association between the number of cycles contributed by each participant and mean menstrual cycle length (data not shown), suggesting the presence of informative cluster size in our data. To address this potential bias in the analysis of experiencing a long and short cycles, we used within-cluster resampling approach with logistic regression models [4].

The details of within-cluster resampling has been published elsewhere [4]. Briefly, we first randomly sampled one cycle per participant with replacement from the original menstrual cycle data. In this data sample, each participant contributed only one observation and the observations are no longer correlated. Therefore, we can fit a logistic regression model for the binary outcome (i.e., experiencing a long or a short cycle) and obtain estimates and standard errors for the regression coefficients. Then we repeated the sampling and analysis steps multiple times and recorded the coefficient estimates and standard errors from each iteration. The final estimates of the regression coefficient is the mean of the coefficient estimates across all iterations. The standard error of the final coefficient estimate $SE(\hat{\beta})$ is given by:

$$SE(\hat{\beta}) = \frac{\sum_q^Q (SE_q)^2}{Q} - \frac{(Q-1)}{Q} S^2$$

Where $SE_q$ is the standard error of the coefficient from the $q^{th}$ $(q=1,2,\ldots,Q)$ iteration. $S^2$ is the variance of the coefficient estimates across all $Q$ iterations.

It has been recommended to perform many iterations to ensure the analysis had sufficient resamples, although no recommendations on the minimum number of iterations are given. We repeated the resampling procedure for $Q$=1,000, 5,000, 7,500, and 10,000 iterations in preliminary analysis and the estimates and standard errors were similar (data not shown). Therefore, we presented results using 10,000 iterations. The ORs and 95%CIs for experiencing a long and short cycle estimated by within-cluster resampling were similar to those estimated using logistic regression with GEE (data not shown).

**Table SM1 Distributions of median cycle length and median cycle length difference before and after excluding artifacts, and threshold values among 49,238 participants under age 50 of AWHS**

| | | Percentiles | | | |
|---|---|---|---|---|---|
| | P10 | P25 | P50 | P75 | P90 |
| Median cycle length | | | | | |
| Before | 25 | 27 | 28·5 | 31·5 | 37 |
| After | 25 | 26·5 | 28 | 30 | 33 |
| Median cycle length difference | | | | | |
| Before | 1 | 2 | 3 | 7 | 16 |
| After | 1 | 2 | 3 | 6 | 14 |
| Threshold value for artifacts | 43 | 44 | 47 | 53 | 67 |

Abbreviation: AWHS, Apple Women's Health Study

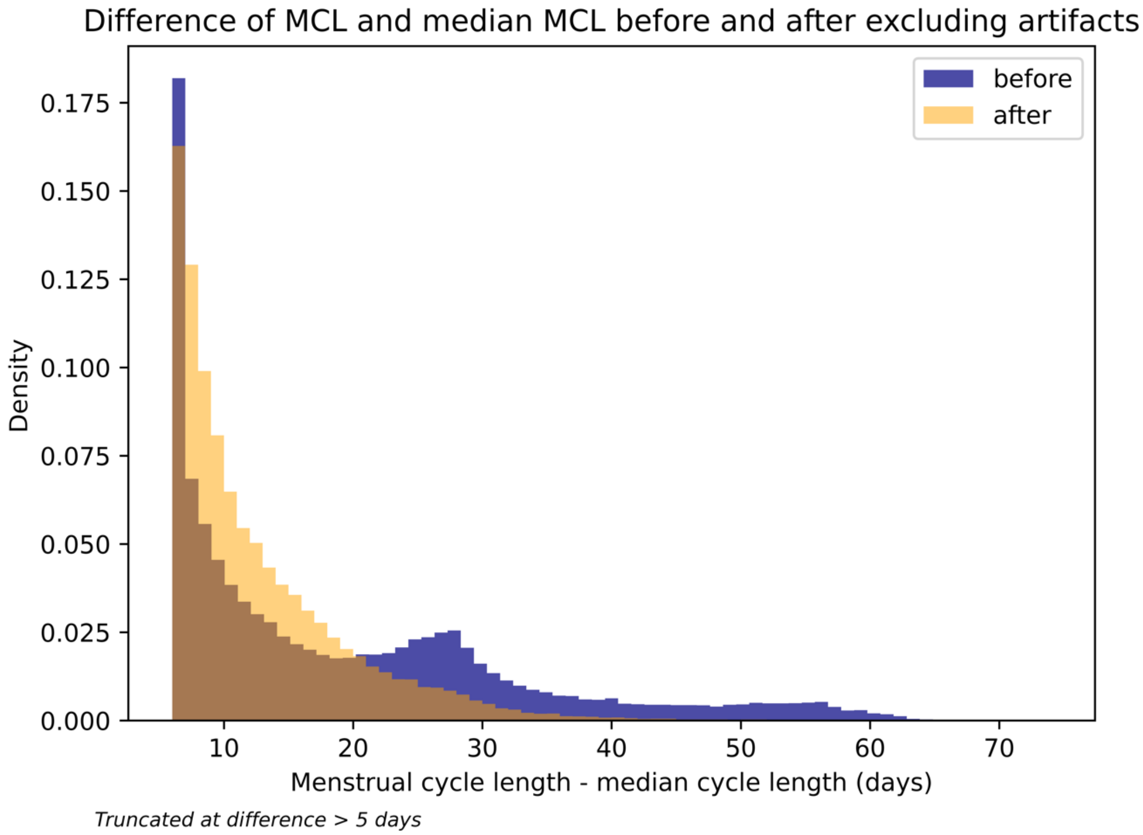


**Figure SM1 Histogram of the difference between the length of each menstrual cycle to the median cycle length of that individual before and after excluding cycle artifacts.**
Started from difference of 5 days.

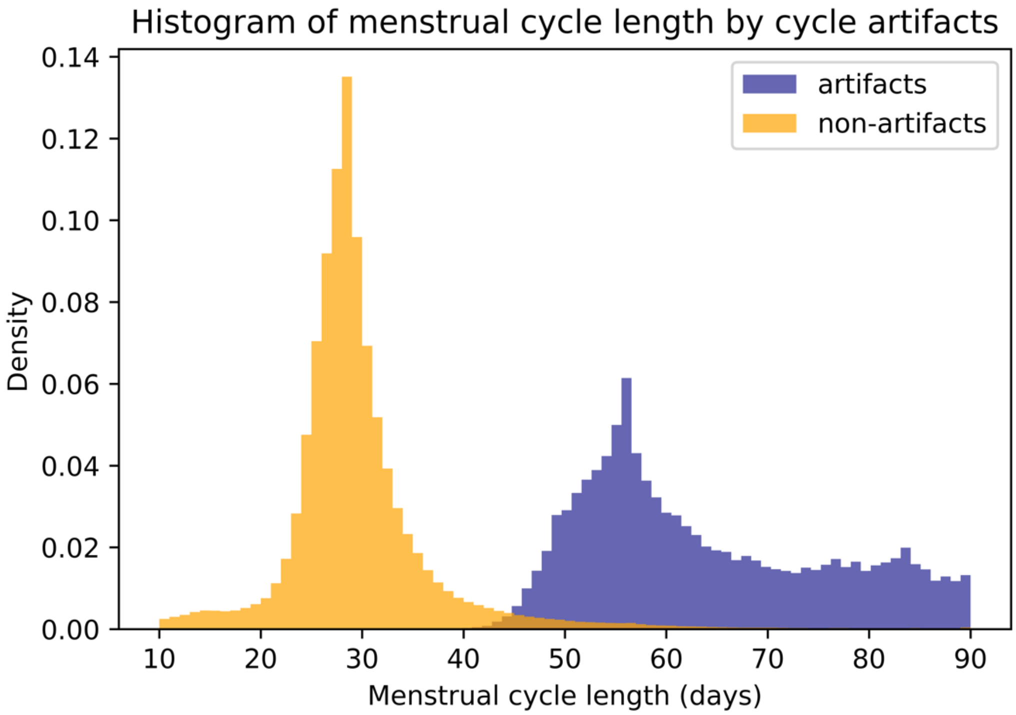


**Figure SM2 Histograms of cycle length among cycles that were identified as potential artifacts and those that were not.**

**Table S1 Number of menstrual cycles by age and BMI across race/ethnicity groups in 165,668 menstrual cycles from 12,608 participants of AWHS**

| | Race/ethnicity | | | | | |
|---|---|---|---|---|---|---|
| | White (N=117,951) | Black (N=9,044) | Asian (N=7,856) | Hispanic (N=11,652) | Other [a] (N=3,658) | More than one (N=15,867) |
| Age | | | | | | |
| under 20 | 5,865 | 627 | 504 | 1,129 | 244 | 1,366 |
| 20-24 | 14,142 | 1,307 | 1,541 | 1,916 | 635 | 2,952 |
| 25-29 | 17,159 | 1,429 | 1,708 | 2,062 | 680 | 2,714 |
| 30-34 | 20,900 | 1,475 | 1,179 | 2,255 | 798 | 2,431 |
| 35-39 | 23,706 | 1,705 | 1,128 | 1,989 | 474 | 2,791 |
| 40-44 | 20,093 | 1,544 | 1,004 | 1,544 | 482 | 2,018 |
| 45-49 | 12,144 | 781 | 604 | 709 | 241 | 1,303 |
| above 50 | 3,582 | 176 | 188 | 48 | 104 | 292 |
| BMI [b] | | | | | | |
| Underweight | 2,907 | 150 | 586 | 358 | 49 | 407 |
| Healthy | 45,357 | 2,090 | 4,640 | 3,592 | 1,503 | 5,565 |
| Overweight | 29,730 | 2,286 | 1,764 | 3,257 | 1,027 | 3,931 |
| Class 1 obese | 19,243 | 1,819 | 644 | 2,471 | 665 | 3,189 |
| Class 2 obese | 10,917 | 1,346 | 216 | 1,144 | 274 | 1,398 |
| Class 3 obese | 9,437 | 1,353 | 6 | 830 | 140 | 1,377 |

Abbreviations: AWHS, Apple Women's Health Study; BMI, body mass index.

[a] Including American Indian or Alaska Native, Middle Eastern or North African, Native Hawaiian or Pacific Islander, or other unspecified race/ethnicity.

[b] Underweight: $BMI<18{\cdot}5\ kg/m^2$; healthy: $18{\cdot}5\leq BMI<25\ kg/m^2$, overweight: $25\leq BMI<30\ kg/m^2$, Class 1 obese: $30\leq BMI<35 kg/m^2$, Class 2 obese: $35\leq BMI<40\ kg/m^2$, Class 3 obese: $BMI\geq 40\ kg/m^2$.

**Table S2 Numbers of menstrual cycles across different cycle length groups by age, race/ethnicity, and BMI in 165,668 menstrual cycles from 12,608 participants of AWHS**

| | Cycle length | | | | | | |
|---|---|---|---|---|---|---|---|
| | <24 days (N=14,976) | 24-26 days (N=40,663) | 27-29 days (N=57,221) | 30-32 days (N=28,095) | 33-35 days (N=11,419) | 36-38 Days (N=5,141) | >38 days (N=8,153) |
| Age | | | | | | | |
| under 20 | 751 | 1,433 | 2,915 | 2,057 | 1,167 | 601 | 811 |
| 20-24 | 1,623 | 3,433 | 7,436 | 5,024 | 2,340 | 1,090 | 1,547 |
| 25-29 | 1,562 | 4,595 | 9,130 | 5,492 | 2,343 | 1,044 | 1,586 |
| 30-34 | 1,949 | 6,801 | 10,508 | 5,458 | 2,113 | 906 | 1,303 |
| 35-39 | 2,581 | 9,289 | 11,974 | 4,696 | 1,656 | 667 | 930 |
| 40-44 | 3,174 | 8,876 | 9,287 | 3,240 | 1,027 | 420 | 661 |
| 45-49 | 2,517 | 5,152 | 4,935 | 1,650 | 576 | 287 | 665 |
| above 50 | 819 | 1,084 | 1,036 | 478 | 197 | 126 | 650 |
| Race/ethnicity | | | | | | | |
| White | 11,264 | 29,955 | 40,745 | 19,175 | 7,655 | 3,330 | 5,467 |
| Black | 806 | 2,621 | 3,142 | 1,323 | 511 | 246 | 395 |
| Asian | 517 | 1,364 | 2,558 | 1,667 | 774 | 420 | 556 |
| Hispanic | 737 | 2,335 | 4,095 | 2,248 | 984 | 487 | 766 |
| Other [a] | 322 | 936 | 1,235 | 662 | 252 | 95 | 156 |
| More than one race/ethnicity | 1,330 | 3,452 | 5,446 | 3,020 | 1,243 | 563 | 813 |
| BMI ($kg/m^2$) [b] | | | | | | | |
| Underweight | 393 | 926 | 1,457 | 855 | 379 | 169 | 278 |
| Healthy | 5,892 | 15,332 | 22,096 | 10,736 | 4,150 | 1,834 | 2,707 |
| Overweight | 3,778 | 10,500 | 14,824 | 7,043 | 2,673 | 1,231 | 1,946 |
| Class 1 obese | 2,613 | 7,130 | 9,412 | 4,618 | 1,929 | 860 | 1,469 |
| Class 2 obese | 1,228 | 3,724 | 5,292 | 2,546 | 1,177 | 518 | 810 |
| Class 3 obese | 1,072 | 3,051 | 4,140 | 2,297 | 1,111 | 529 | 943 |

Abbreviations: AWHS, Apple Women's Health Study; BMI, body mass index.

[a] Including American Indian or Alaska Native, Middle Eastern or North African, Native Hawaiian or Pacific Islander, or other unspecified race/ethnicity.

[b] Underweight: BMI<18·5 $kg/m^2$; healthy: 18·5≤BMI<25 $kg/m^2$, overweight: 25≤BMI<30 $kg/m^2$, Class 1 obese: 30≤BMI<35$kg/m^2$, Class 2 obese: 35≤BMI<40 $kg/m^2$, Class 3 obese: BMI≥40 $kg/m^2$.

**Table S3 Differences and 95% confidence intervals (95%CIs) of mean menstrual cycle length with age, race/ethnicity, and BMI after restricting to participants who tracked ≥ 3 cycles, to participants under age 50, to cycles with confirmed accurate tracking, to cycles with complete data, and including participants with uterine fibroids**

| | Restricted to participants with ≥ 3 cycles [a] | Restricted to participants under age 50 [a] | Restricted to cycles with accurate tracking [a] | Restricted to cycles with complete data [a] | Including participants with uterine fibroids [a] |
|---|---|---|---|---|---|
| N cycles | 163,261 | 161,278 | 54,804 | 150,197 | 175,703 |
| **Age** | | | | | |
| under 20 | 1·59 (1·29, 1·90) | 1·58 (1·27, 1·89) | 1·79 (1·25, 2·34) | 1·47 (1·14, 1·81) | 1·67 (1·36, 1·99) |
| 20-24 | 1·39 (1·15, 1·63) | 1·40 (1·16, 1·63) | 1·71 (1·36, 2·05) | 1·43 (1·18, 1·68) | 1·47 (1·23, 1·70) |
| 25-29 | 1·09 (0·89, 1·29) | 1·09 (0·89, 1·29) | 1·57 (1·27, 1·87) | 1·13 (0·92, 1·35) | 1·13 (0·93, 1·33) |
| 30-34 | 0·55 (0·38, 0·71) | 0·55 (0·39, 0·71) | 0·71 (0·45, 0·98) | 0·56 (0·38, 0·73) | 0·58 (0·42, 0·75) |
| 35-39 | Reference | Reference | Reference | Reference | Reference |
| 40-44 | -0·47 (-0·64, -0·31) | -0·51 (-0·67, -0·35) | -0·54 (-0·81, -0·27) | -0·43 (-0·61, -0·26) | -0·49 (-0·66, -0·33) |
| 45-49 | -0·31 (-0·53, -0·08) | -0·48 (-0·71, -0·25) | -0·44 (-0·77, -0·10) | -0·29 (-0·53, -0·05) | -0·30 (-0·52, -0·09) |
| above 50 | 1·94 (1·57, 2·31) | - | 2·02 (1·49, 2·54) | 2·02 (1·63, 2·40) | 2·00 (1·65, 2·34) |
| **Race/ethnicity** | | | | | |
| White | Reference | Reference | Reference | Reference | Reference |
| Black | -0·30 (-0·67, 0·06) | -0·26 (-0·64, 0·12) | -0·56 (-1·01, -0·11) | -0·38 (-0·78, 0·01) | -0·20 (-0·54, 0·15) |
| Asian | 1·62 (1·23, 2·01) | 1·62 (1·21, 2·04) | 1·19 (0·71, 1·67) | 1·52 (1·09, 1·95) | 1·49 (1·09, 1·89) |
| Hispanic | 0·80 (0·49, 1·11) | 0·73 (0·41, 1·05) | 0·67 (0·28, 1·06) | 0·72 (0·39, 1·05) | 0·77 (0·46, 1·07) |
| Other [b] | -0·03 (-0·57, 0·51) | 0·30 (-0·26, 0·86) | -0·42 (-1·08, 0·24) | -0·09 (-0·69, 0·51) | 0·25 (-0·29, 0·79) |
| More than one race/ethnicity | 0·19 (-0·08, 0·46) | 0·16 (-0·12, 0·44) | 0·26 (-0·06, 0·58) | 0·04 (-0·25, 0·33) | 0·15 (-0·12, 0·43) |
| **BMI [c]** | | | | | |
| Underweight | 0·11 (-0·35, 0·57) | 0·02 (-0·45, 0·50) | 0·94 (0·28, 1·59) | 0·17 (-0·33, 0·67) | 0·03 (-0·44, 0·51) |
| Healthy | Reference | Reference | Reference | Reference | Reference |
| Overweight | 0·27 (0·09, 0·46) | 0·22 (0·03, 0·41) | 0·17 (-0·07, 0·41) | 0·29 (0·10, 0·49) | 0·23 (0·04, 0·41) |
| Class 1 obese | 0·51 (0·29, 0·73) | 0·57 (0·34, 0·79) | 0·55 (0·27, 0·83) | 0·54 (0·31, 0·78) | 0·57 (0·35, 0·79) |
| Class 2 obese | 0·77 (0·50, 1·04) | 0·75 (0·48, 1·03) | 0·77 (0·43, 1·12) | 0·74 (0·45, 1·03) | 0·76 (0·49, 1·03) |
| Class 3 obese | 1·50 (1·20, 1·79) | 1·49 (1·18, 1·79) | 1·51 (1·13, 1·89) | 1·59 (1·28, 1·91) | 1·57 (1·27, 1·87) |

Abbreviations: BMI, body mass index.

[a] Exclusively adjusting for age, race/ethnicity, body weight, and additionally for smoking, alcohol drinking, parity, physical activity, education, perceived stress scores, and MacArthur scale of subjective social status.
[b] Including American Indian or Alaska Native, Middle Eastern or North African, Native Hawaiian or Pacific Islander, or other unspecified race/ethnicity.
[c] Underweight: BMI<18·5 kg/$m^2$; healthy: 18·5≤BMI<25 kg/$m^2$, overweight: 25≤BMI<30 kg/$m^2$, class 1 obese: 30 ≤BMI<35 kg/$m^2$, class 2 obese: 35≤BMI<40 kg/$m^2$, class 3 obese: BMI≥40 kg/$m^2$.

**Table S4 Differences and 95% confidence intervals (95%CIs) of median menstrual cycle length with age, race/ethnicity, and BMI to participants who tracked ≥ 3 cycles, to participants under age 50, to cycles with confirmed accurate tracking, to cycles with complete data, and including participants with uterine fibroids**

| | Restricted to participants with ≥ 3 cycles [a] | Restricted to participants under age 50 [a] | Restricted to cycles with accurate tracking [a] | Restricted to cycles with complete data [a] | Including participants with uterine fibroids [a] |
|---|---|---|---|---|---|
| N cycles | 163,261 | 161,278 | 54,804 | 150,197 | 175,703 |
| **Age** | | | | | |
| under 20 | 1·98 (1·42, 2·54) | 1·97 (1·56, 2·37) | 1·87 (1·33, 2·41) | 1·89 (1·32, 2·46) | 1·93 (1·46, 2·40) |
| 20-24 | 1·54 (1·01,2·07) | 1·49 (1·05, 1·94) | 1·72 (1·41, 2·03) | 1·57 (1·01, 2·12) | 1·56 (1·12, 2·00) |
| 25-29 | 1·08 (0·73, 1·42) | 1·00 (0·73, 1·27) | 1·36 (1·01, 1·70) | 1·13 (0·80 1·45) | 1·27 (1·00, 1·53) |
| 30-34 | 0·54 (0·28, 0·80) | 0·49 (0·23, 0·76) | 0·74 (0·47, 1·01) | 0·57 (0·33, 0·80) | 0·71 (0·50, 0·91) |
| 35-39 | Reference | Reference | Reference | Reference | Reference |
| 40-44 | -0·46 (-0·66, -0·26) | -0·51 (-0·70, -0·31) | -0·56 (-0·92, 0·20) | -0·43 (-0·63, -0·23) | -0·37 (-0·54, -0·19) |
| 45-49 | -0·46 (-0·72, -0·21) | -0·51 (-0·75, -0·26) | -0·53 (-0·88, -0·18) | -0·43 (-0·70, -0·17) | -0·37 (-0·57, -0·16) |
| above 50 | 0·55 (-0·22, 1·33) | - | 0·22 (-0·41, 0·85) | 1·01 (0·38, 1·64) | 0·77 (0·18, 1·35) |
| **Race/ethnicity** | | | | | |
| White | Reference | Reference | Reference | Reference | Reference |
| Black | 0·00 (-0·57, 0·57) | 0·00 (-0·49, 0·49) | -0·58 (-1·00, -0·16) | -0·12 (-0·69, 0·46) | 0·00 (-0·45, 0·45) |
| Asian | 1·38 (0·77, 1·98) | 1·48 (1·01, 1·95) | 1·07 (0·59, 1·56) | 1·12 (0·54, 1·69) | 1·22 (0·83, 1·61) |
| Hispanic | 0·54 (-0·05, 1·13) | 1·00 (0·43, 1·57) | 0·55 (0·17, 0·93) | 0·66(0·08, 1·24) | 0·78 (0·22, 1·34) |
| Other [b] | -0·46 (-0·95, 0·03) | -0·49 (-1·04, -0·05) | -0·46 (-1·01, 0·08) | -0·45 (-0·95, 0·05) | -0·30 (-0·85, 0·25) |
| More than one race/ethnicity | 0·00 (-0·43, 0·43) | 0·00 (-0·29, 0·29) | 0·22 (-0·08, 0·52) | 0·00 (-0·35 0·35) | 0·07 (-0·21, 0·35) |
| **BMI [c]** | | | | | |
| Underweight | 0·00 (-0·46, 0·46) | -0·01 (-0·45, 0·43) | 0·90 (0·37, 1·43) | 0·29 (-0·34, 0·93) | 0·08 (-0·39, 0·54) |
| Healthy | Reference | Reference | Reference | Reference | Reference |
| Overweight | 0·46 (0·07, 0·86) | 0·49 (0·21, 0·77 | 0·27 (-0·02, 0·56) | 0·44 (0·08, 0·79) | 0·22 (-0·05, 0·49) |
| Class 1 obese | 0·46 (0·04, 0·88) | 0·49 (0·11, 0·88) | 0·32 (0·07, 0·57) | 0·44 (0·02, 0·86) | 0·29 (-0·04, 0·62) |
| Class 2 obese | 0·53 (0·10, 0·95) | 0·99 (0·56, 1·42) | 0·58 (0·25, 0·92) | 0·57 (0·16, 0·97) | 0·66 (0·33, 0·99) |
| Class 3 obese | 1·46 (1·03, 1·89) | 1·49 (1·13, 1·86) | 1·15 (0·65, 1·65) | 1·44 (1·01, 1·87) | 1·29 (1·01, 1·58) |

Abbreviations: BMI, body mass index.

[a] Exclusively adjusting for age, race/ethnicity, body weight, and additionally for smoking, alcohol drinking, parity, physical activity, education, perceived stress scores, and MacArthur scale of subjective social status.
[b] Including American Indian or Alaska Native, Middle Eastern or North African, Native Hawaiian or Pacific Islander, or other unspecified race/ethnicity.
[c] Underweight: BMI<18·5 kg/$m^2$; healthy: 18·5≤BMI<25 kg/$m^2$, overweight: 25≤BMI<30 kg/$m^2$, class 1 obese: 30 ≤BMI<35 kg/$m^2$, class 2 obese: 35≤BMI<40 kg/$m^2$, class 3 obese: BMI≥40 kg/$m^2$.

**Table S5 Odds ratios (ORs) and 95% confidence intervals (95%CIs) of experiencing a short (<24 days) menstrual cycle by age, race/ethnicity, and BMI to participants who tracked ≥ 3 cycles, to participants under age 50, to cycles with confirmed accurate tracking, to cycles with complete data, and including participants with uterine fibroids**

| | Restricted to participants with ≥ 3 cycles [a] | Restricted to participants under age 50 [a] | Restricted to cycles with accurate tracking [a] | Restricted to cycles with complete data [a] | Including participants with uterine fibroids [a] |
|---|---|---|---|---|---|
| N short cycles/N total [b] | 14,796/155,433 | 14,157/153,775 | 5,171/52,593 | 14,976/157,515 | 16,366/167,127 |
| Age | | | | | |
| under 20 | 0·96 (0·79, 1·16) | 0·90 (0·73, 1·10) | 0·79 (0·56, 1·11) | 0·93 (0·75, 1·15) | 0·87 (0·72, 1·06) |
| 20-24 | 0·97 (0·84, 1·12) | 0·91 (0·79, 1·06) | 0·78 (0·63, 0·97) | 0·86 (0·73, 1·01) | 0·90 (0·78, 1·04) |
| 25-29 | 0·75 (0·65, 0·85) | 0·77 (0·67, 0·88) | 0·72 (0·59, 0·87) | 0·74 (0·64, 0·86) | 0·77 (0·67, 0·88) |
| 30-34 | 0·83 (0·73, 0·93) | 0·83 (0·73, 0·94) | 0·84 (0·71, 1·00) | 0·82 (0·72, 0·94) | 0·83 (0·74, 0·93) |
| 35-39 | Reference | Reference | Reference | Reference | Reference |
| 40-44 | 1·52 (1·37, 1·70) | 1·49 (1·34, 1·67) | 1·48 (1·27, 1·71) | 1·50 (1·33, 1·69) | 1·51 (1·36, 1·68) |
| 45-49 | 2·45 (2·18, 2·75) | 2·46 (2·19, 2·77) | 2·47 (2·10, 2·90) | 2·40 (2·12, 2·73) | 2·40 (2·14, 2·68) |
| above 50 | 3·33 (2·83, 3·91) | - | 3·43 (2·76, 4·27) | 3·33 (2·79, 3·97) | 3·31 (2·84, 3·85) |
| Race/ethnicity | | | | | |
| White | Reference | Reference | Reference | Reference | Reference |
| Black | 0·97 (0·83, 1·13) | 0·98 (0·82, 1·16) | 1·05 (0·83, 1·33) | 0·99 (0·83, 1·17) | 0·97 (0·84, 1·12) |
| Asian | 0·69 (0·56, 0·85) | 0·67 (0·54, 0·84) | 0·82 (0·62, 1·09) | 0·67 (0·53, 0·85) | 0·68 (0·56, 0·84) |
| Hispanic | 0·79 (0·68, 0·92) | 0·87 (0·75, 1·03) | 0·79 (0·63, 1·00) | 0·87 (0·73, 1·04) | 0·87 (0·75, 1·01) |
| Other [c] | 0·98 (0·77, 1·25) | 0·85 (0·66, 1·10) | 0·89 (0·63, 1·27) | 0·94 (0·71, 1·23) | 0·86 (0·68, 1·10) |
| More than one race/ethnicity | 0·95 (0·84, 1·08) | 1·00 (0·88, 1·15) | 1·03 (0·87, 1·23) | 1·06 (0·93, 1·22) | 1·01 (0·89, 1·15) |
| BMI [d] | | | | | |
| Underweight | 1·00 (0·82, 1·22) | 0·99 (0·80, 1·22) | 1·07 (0·77, 1·48) | 0·89 (0·71, 1·13) | 0·97 (0·78, 1·20) |
| Healthy | Reference | Reference | Reference | Reference | Reference |
| Overweight | 0·90 (0·82, 0·99) | 0·91 (0·83, 1·00) | 0·94 (0·83, 1·06) | 0·90 (0·81, 0·99) | 0·94 (0·86, 1·03) |
| Class 1 obese | 0·91 (0·82, 1·01) | 0·95 (0·85, 1·05) | 0·97 (0·84, 1·12) | 0·90 (0·81, 1·01) | 0·91 (0·82, 1·01) |
| Class 2 obese | 0·83 (0·73, 0·95) | 0·78 (0·69, 0·90) | 0·82 (0·68, 0·98) | 0·76 (0·66, 0·87) | 0·79 (0·70, 0·90) |
| Class 3 obese | 0·85 (0·73, 0·97) | 0·80 (0·69, 0·93) | 0·86 (0·71, 1·05) | 0·74 (0·64, 0·86) | 0·80 (0·70, 0·91) |

Abbreviations: BMI, body mass index.
[a] Exclusively adjusting for age, race/ethnicity, body weight, season, and region, and additionally for smoking, alcohol drinking, parity, physical activity, education, perceived stress scores, and MacArthur scale of subjective social status.
[b] Long cycles were excluded from the total.
[c] Including American Indian or Alaska Native, Middle Eastern or North African, Native Hawaiian or Pacific Islander, or other unspecified race/ethnicity.
[d] Underweight: BMI<18·5 kg/$m^2$; healthy: 18·5≤BMI<25 kg/$m^2$, overweight: 25≤BMI<30 kg/$m^2$, class 1 obese: 30 ≤BMI<35 kg/$m^2$, class 2 obese: 35≤BMI<40 kg/$m^2$, class 3 obese: BMI≥40 kg/$m^2$.

**Table S6 Odds ratios (ORs) and 95% confidence intervals (95%CIs) of experiencing a long (>38 days) menstrual cycle by age, race/ethnicity, and BMI, after restricting to women who tracked ≥ 3 cycles, to women under age 50, to cycles with confirmed accurate tracking, to cycles with complete data, and including participants with uterine fibroids**

| | Restricted to participants with ≥ 3 cycles [a] | Restricted to participants under age 50 [a] | Restricted to cycles with accurate tracking [a] | Restricted to cycles with complete data [a] | Including participants with uterine fibroids [a] |
|---|---|---|---|---|---|
| N long cycles/N total [b] | 7,828/148,465 | 7,503/153,775 | 2,396/49,818 | 8,153/150,692 | 8,591/159,337 |
| Age | | | | | |
| under 20 | 2·02 (1·61, 2·54) | 1·78 (1·42, 2·24) | 1·95 (1·34, 2·84) | 1·78 (1·40, 2·26) | 1·83 (1·46, 2·29) |
| 20-24 | 2·03 (1·69, 2·44) | 1·83 (1·53, 2·19) | 1·87 (1·44, 2·42) | 1·81 (1·49, 2·20) | 1·85 (1·55, 2·21) |
| 25-29 | 1·76 (1·48, 2·10) | 1·67 (1·41, 1·99) | 1·75 (1·38, 2·22) | 1·69 (1·41, 2·02) | 1·67 (1·41, 1·98) |
| 30-34 | 1·33 (1·12, 1·58) | 1·28 (1·08, 1·52) | 1·32 (1·04, 1·67) | 1·30 (1·08, 1·55) | 1·27 (1·08, 1·51) |
| 35-39 | Reference | Reference | Reference | Reference | Reference |
| 40-44 | 0·99 (0·82, 1·21) | 0·99 (0·82, 1·20) | 0·92 (0·71, 1·19) | 1·04 (0·85, 1·27) | 0·98 (0·82, 1·18) |
| 45-49 | 2·05 (1·68, 2·49) | 1·84 (1·51, 2·23) | 1·68 (1·29, 2·19) | 1·76 (1·43, 2·16) | 1·71 (1·42, 2·05) |
| above 50 | 6·93 (5·64, 8·53) | - | 6·78 (5·10, 9·02) | 6·59 (5·30, 8·19) | 6·44 (5·30, 7·82) |
| Race/ethnicity | | | | | |
| White | Reference | Reference | Reference | Reference | Reference |
| Black | 0·97 (0·79, 1·20) | 1·07 (0·87, 1·33) | 0·87 (0·63, 1·20) | 1·00 (0·80, 1·25) | 1·11 (0·92, 1·35) |
| Asian | 1·51 (1·23, 1·85) | 1·46 (1·18, 1·80) | 1·34 (1·00, 1·81) | 1·42 (1·14, 1·77) | 1·40 (1·15, 1·72) |
| Hispanic | 1·28 (1·10, 1·51) | 1·25 (1·07, 1·46) | 1·33 (1·05, 1·69) | 1·27 (1·07, 1·50) | 1·27 (1·09, 1·49) |
| Other [c] | 0·95 (0·69, 1·30) | 1·08 (0·78, 1·48) | 0·90 (0·56, 1·44) | 0·95 (0·67, 1·34) | 1·08 (0·80, 1·47) |
| More than one race/ethnicity | 0·97 (0·84, 1·14) | 1·01 (0·86, 1·18) | 1·16 (0·93, 1·45) | 1·00 (0·85, 1·18) | 1·01 (0·87, 1·17) |
| BMI [d] | | | | | |
| Underweight | 1·29 (0·99, 1·68) | 1·23 (0·93, 1·62) | 1·96 (1·34, 2·85) | 1·32 (0·98, 1·77) | 1·22 (0·93, 1·60) |
| Healthy | Reference | Reference | Reference | Reference | Reference |
| Overweight | 1·14 (1·01, 1·29) | 1·08 (0·95, 1·22) | 1·05 (0·88, 1·26) | 1·16 (1·02, 1·32) | 1·11 (0·98, 1·24) |
| Class 1 obese | 1·33 (1·16, 1·52) | 1·32 (1·15, 1·52) | 1·38 (1·14, 1·68) | 1·32 (1·14, 1·52) | 1·30 (1·14, 1·49) |
| Class 2 obese | 1·41 (1·20, 1·65) | 1·35 (1·14, 1·60) | 1·48 (1·18, 1·86) | 1·34 (1·13, 1·60) | 1·35 (1·16, 1·59) |
| Class 3 obese | 1·99 (1·69, 2·34) | 1·76 (1·49, 2·08) | 2·07 (1·63, 2·62) | 1·78 (1·50, 2·12) | 1·80 (1·54, 2·11) |

Abbreviations: BMI, body mass index.

[a] Exclusively adjusting for age, race/ethnicity, body weight, and additionally for smoking, alcohol drinking, parity, physical activity, education, perceived stress scores, and MacArthur scale of subjective social status.
[b] Short cycles were excluded from the total.
[c] Including American Indian or Alaska Native, Middle Eastern or North African, Native Hawaiian or Pacific Islander, or other unspecified race/ethnicity.
[d] Underweight: BMI<18·5 kg/$m^2$; healthy: 18·5≤BMI<25 kg/$m^2$, overweight: 25≤BMI<30 kg/$m^2$, class 1 obese: 30 ≤BMI<35 kg/$m^2$, class 2 obese: 35≤BMI<40 kg/$m^2$, class 3 obese: BMI≥40 kg/$m^2$.

**Table S7 Differences and 95% confidence intervals (95%CIs) of mean menstrual cycle length with age, race/ethnicity, and BMI in 59,431 cycles from 4,119 AWHS participants who never had known COVID-19 infection**

| | Participants, N (%) | Cycles, N (%) | Differences and 95%CIs of mean menstrual cycle length (days) | Differences and 95%CIs of median menstrual cycle length (days) |
|---|---|---|---|---|
| Age (years) | | | | |
| under 20 | 166 (4·0) | 2,407 (4·0) | 1·30 (0·71, 1·89) | 1·54 (0·78, 2·29) |
| 20-24 | 458 (11·1) | 6,390 (10·8) | 1·37 (0·95, 1·79) | 1·53 (0·94, 2·12) |
| 25-29 | 688 (16·7) | 8,613 (14·5) | 1·23 (0·88, 1·59) | 1·32 (0·92, 1·73) |
| 30-34 | 786 (19·1) | 10,446 (17·6) | 0·75 (0·46, 1·04) | 0·85 (0·55, 1·16) |
| 35-39 | 774 (18·8) | 11,301 (19·0) | Reference | Reference |
| 40-44 | 637 (15·5) | 10,634 (17·9) | -0·51 (-0·78, -0·24) | -0·44 (-0·65, -0·22) |
| 45-49 | 454 (11·0) | 7,564 (12·7) | -0·40 (-0·76, -0·04) | -0·40 (-0·77· -0·02) |
| above 50 | 156 (3·8) | 2,076 (3·5) | 2·37 (1·80, 2·93) | 0·80 (-0·09, 1·70) |
| Race/ethnicity | | | | |
| White | 3,036 (73·7) | 43,759 (73·6) | Reference | Reference |
| Black | 207 (5·0) | 3,112 (5·2) | -0·04 (-0·69, 0·61) | -0·14 (-0·88, 0·60) |
| Asian | 192 (4·7) | 3,033 (5·1) | 1·41 (0·74, 2·08) | 1·42 (0·70, 2·13) |
| Hispanic | 247 (6·0) | 3,493 (5·9) | 0·84 (0·25, 1·43) | 1·14 (0·32, 1·97) |
| Other [a] | 87 (2·1) | 1,274 (2·1) | 0·25 (-0·70, 1·21) | 0·14 (-0·66, 0·94) |
| More than one race/ethnicity | 350 (8·5) | 4,760 (8·0) | 0·11 (-0·39, 0·62) | -0·14 (-0·60, 0·31) |
| BMI (kg/m$^2$) [b] | | | | |
| Underweight | 99 (2·4) | 1,508 (2·5) | 0·46 (-0·33, 1·25) | 0·57 (-0·10, 1·24) |
| Healthy | 1,541 (37·4) | 22,691 (38·2) | Reference | |
| Overweight | 1,075 (26·1) | 15,445 (26·0) | 0·23 (-0·08, 0·54) | 0·19 (-0·24, 0·62) |
| Class 1 obese | 700 (17·0) | 9,804 (16·5) | 0·62 (0·25, 0·99) | 0·45 (0·03, 0·87) |
| Class 2 obese | 389 (9·4) | 5,422 (9·1) | 0·84 (0·38, 1·31) | 1·16 (0·57, 1·75) |
| Class 3 obese | 315 (7·6) | 4,561 (7·7) | 1·64 (1·11, 2·16) | 1·28 (0·87, 1·69) |

Abbreviations: AWHS, Apple Women's Health Study; BMI, body mass index.

Participants who never had known COVID-19 infection were identified by self-reported not having been previous tested positive for COVID-19 in 2022.

All models were exclusively adjusting for age, race/ethnicity, and BMI, and additionally for smoking, alcohol use, parity, physical activity, education, perceived stress scores, and MacArthur scale of subjective social status. Missing values in age, race/ethnicity, and BMI were excluded. Missing values in other covariates were treated with missing indicator.

[a] Including American Indian or Alaska Native, Middle Eastern or North African, Native Hawaiian or other Pacific Islander, or other unspecified race/ethnicity.

[b] Underweight: BMI<18·5 kg/m$^2$; healthy: 18·5≤BMI<25 kg/m$^2$, overweight: 25≤BMI<30 kg/m$^2$, class 1 obese: 30≤BMI<35 kg/m$^2$, class 2 obese: 35≤BMI<40 kg/m$^2$, class 3 obese: BMI≥40 kg/m$^2$.

**Table S8 Odds ratios (ORs) and 95% confidence intervals (95%CIs) of experiencing menstrual irregularity by age, race/ethnicity, and BMI using different criteria among 11,040 participants with ≥ 3 menstrual cycles in AWHS**

| | Standard deviation of cycle length ≥7 days [a] | Median cycle length difference ≥9 days [a] | Longest and shortest cycle differ by ≥7 days [a] |
|---|---|---|---|
| Age | | | |
| under 20 | 2·83 (2·60, 3·08) | 2·06 (1·82, 2·33) | 3·36 (3·11, 3·63) |
| 20-24 | 3·00 (2·80, 3·22) | 2·47 (2·24, 2·74) | 1·74 (1·65, 1·83) |
| 25-29 | 1·97 (1·84, 2·11) | 2·05 (1·86, 2·25) | 1·46 (1·39, 1·52) |
| 30-34 | 1·56 (1·46, 1·67) | 1·52 (1·38, 1·68) | 1·12 (1·07, 1·16) |
| 35-39 | Reference | Reference | Reference |
| 40-44 | 1·43 (1·34, 1·54) | 1·41 (1·27, 1·56) | 1·25 (1·20, 1·30) |
| 45-49 | 5·11 (4·78, 5·47) | 4·54 (4·13, 5·00) | 2·52 (2·37, 2·67) |
| above 50 | 37·80 (34·50, 41·44) | 22·09 (19·81, 24·63) | 5·75 (4·96, 6·71) |
| Race/ethnicity | | | |
| White | Reference | Reference | Reference |
| Black | 0·91 (0·84, 0·98) | 0·73 (0·65, 0·82) | 0·78 (0·74, 0·83) |
| Asian | 1·27 (1·18, 1·38) | 1·08 (0·96, 1·20) | 1·18 (1·11, 1·26) |
| Hispanic | 1·41 (1·33, 1·50) | 1·39 (1·28, 1·51) | 1·06 (1·00, 1·12) |
| Other [b] | 0·67 (0·58, 0·75) | 0·78 (0·66, 0·92) | 0·93 (0·85, 1·01) |
| More than one race/ethnicity | 0·86 (0·81, 0·91) | 0·84 (0·77, 0·91) | 0·99 (0·94, 1·03) |
| BMI [c] | | | |
| Underweight | 1·40 (1·27, 1·53) | 1·19 (1·03, 1·37) | 1·22 (1·12, 1·34) |
| Healthy | Reference | Reference | Reference |
| Overweight | 1·06 (1·01, 1·10) | 1·26 (1·19, 1·34) | 1·03 (1·00, 1·07) |
| Class 1 obese | 1·24 (1·18, 1·30) | 1·36 (1·27, 1·46) | 1·07 (1·03, 1·11) |
| Class 2 obese | 1·29 (1·21, 1·37) | 1·41 (1·30, 1·54) | 1·16 (1·10, 1·22) |
| Class 3 obese | 1·71 (1·61, 1·83) | 1·91 (1·76, 2·08) | 1·41 (1·34, 1·49) |

[a] Exclusively adjusting for age, race/ethnicity, body weight, and additionally for smoking, alcohol drinking, parity, physical activity, education, perceived stress scores, and MacArthur scale of subjective social status.

[b] Including American Indian or Alaska Native, Middle Eastern or North African, Native Hawaiian or Pacific Islander, or other unspecified race/ethnicity.
[c] Underweight: BMI<18·5 kg/m$^2$; healthy: 18·5≤BMI<25 kg/m$^2$, overweight: 25≤BMI<30 kg/m$^2$, class 1 obese: 30 ≤BMI<35 kg/m$^2$, class 2 obese: 35≤BMI<40 kg/m$^2$, class 3 obese: BMI≥40 kg/m$^2$.

**Table S9 Differences and 95% confidence intervals of mean menstrual cycle length by BMI groups using categorical, continuous, and piecewise BMI**

| | Median BMI ($kg/m^2$) | Difference in mean menstrual cycle length | | |
|---|---|---|---|---|
| | | Main model (Categorical BMI) | Continuous BMI | Piecewise BMI[a] |
| Underweight | 17·8 | 0·04 (-0·44, 0·51) | 0·27 (0·18, 0·36) | 0·04 (-14·5, 14·5) |
| Healthy | 22·3 | Reference | Reference | Reference |
| Overweight | 27·3 | 0·26 (0·07, 0·45) | 0·27 (0·18, 0·36) | 0·27 (0·18, 0·36) |
| Obese 1 | 32·1 | 0·54 (0·32, 0·77) | 0·56 (0·38, 0·74) | 0·56 (0·38, 0·74) |
| Obese 2 | 37·2 | 0·76 (0·48, 1·03) | 0·86 (0·58, 1·14) | 0·86 (0·58, 1·14) |
| Obese 3 | 44·1 | 1·54 (1·24, 1·85) | 1·28 (0·86, 1·70) | 1·28 (0·86, 1·70) |

Abbreviations: BMI, body mass index.

All models were adjusted for age, race/ethnicity, additionally for smoking, alcohol use, parity, physical activity, education, perceived stress scores, and MacArthur scale of subjective social status. The difference and 95% confidence intervals by BMI categories in the continuous and piecewise BMI models were calculated using the differences of the median BMI in each category and the one in the healthy BMI category.

Underweight: BMI<18·5 $kg/m^2$; healthy: 18·5≤BMI<25 $kg/m^2$, overweight: 25≤BMI<30 $kg/m^2$, class 1 obese: 30≤BMI<35 $kg/m^2$, class 2 obese: 35≤BMI<40 $kg/m^2$, class 3 obese: BMI≥40 $kg/m^2$.

[a] Piecewise BMI model was fitted by adding a multiplicative interaction term of being underweight (binary) and the continuous BMI in the model with all other covariates remained the same. We considered this model because the main model with categorical BMI suggested potential non-linear relationship of BMI and mean menstrual cycle length.

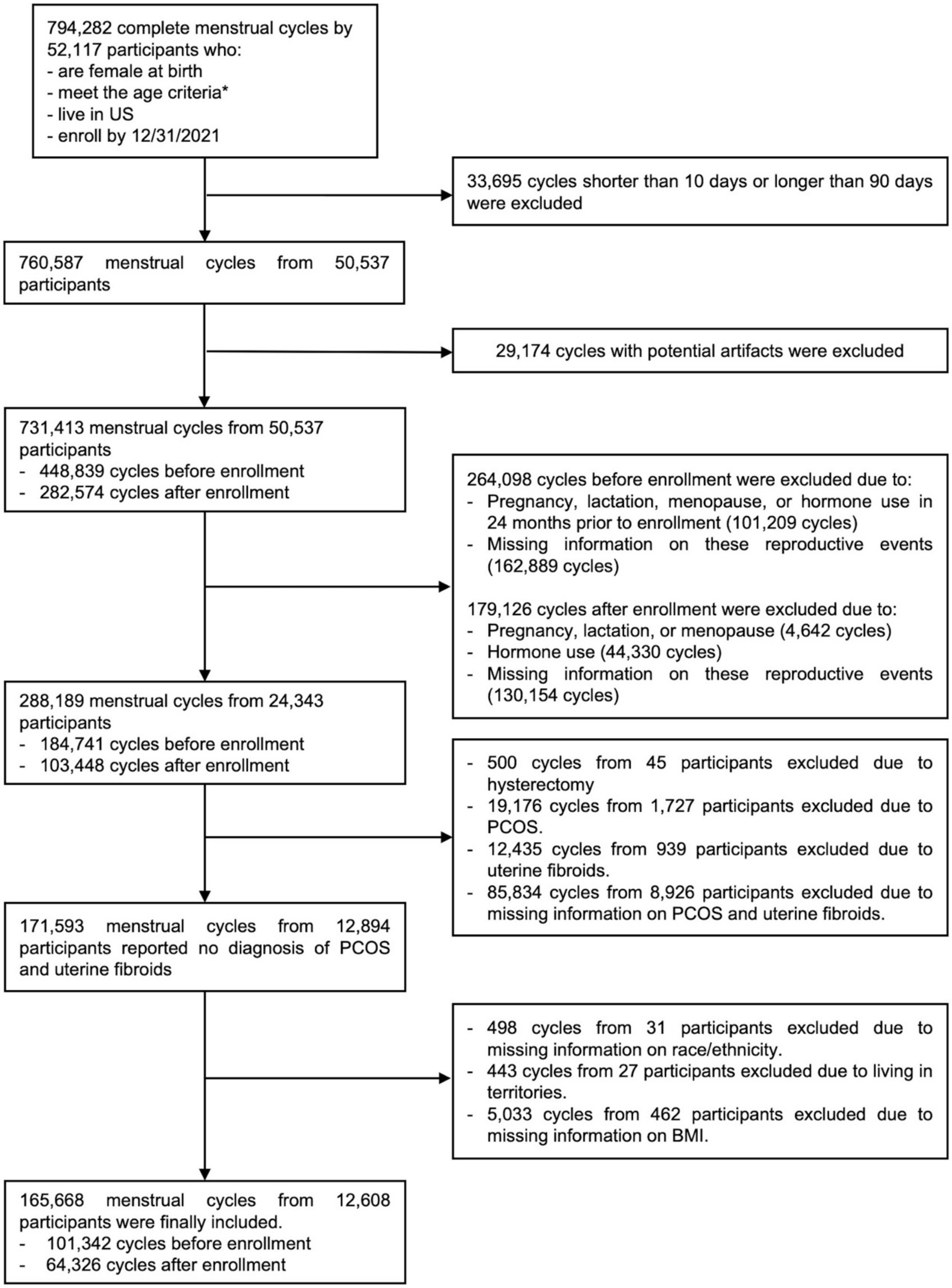


**Figure S1 Inclusion and exclusion of menstrual cycles and AWHS participants**

*Above age 18 for most states, above age 19 for Alabama and Nebraska, above 21 for Puerto Rico

Abbreviations: AWHS, Apple Women's Health Study; BMI, body mass index.

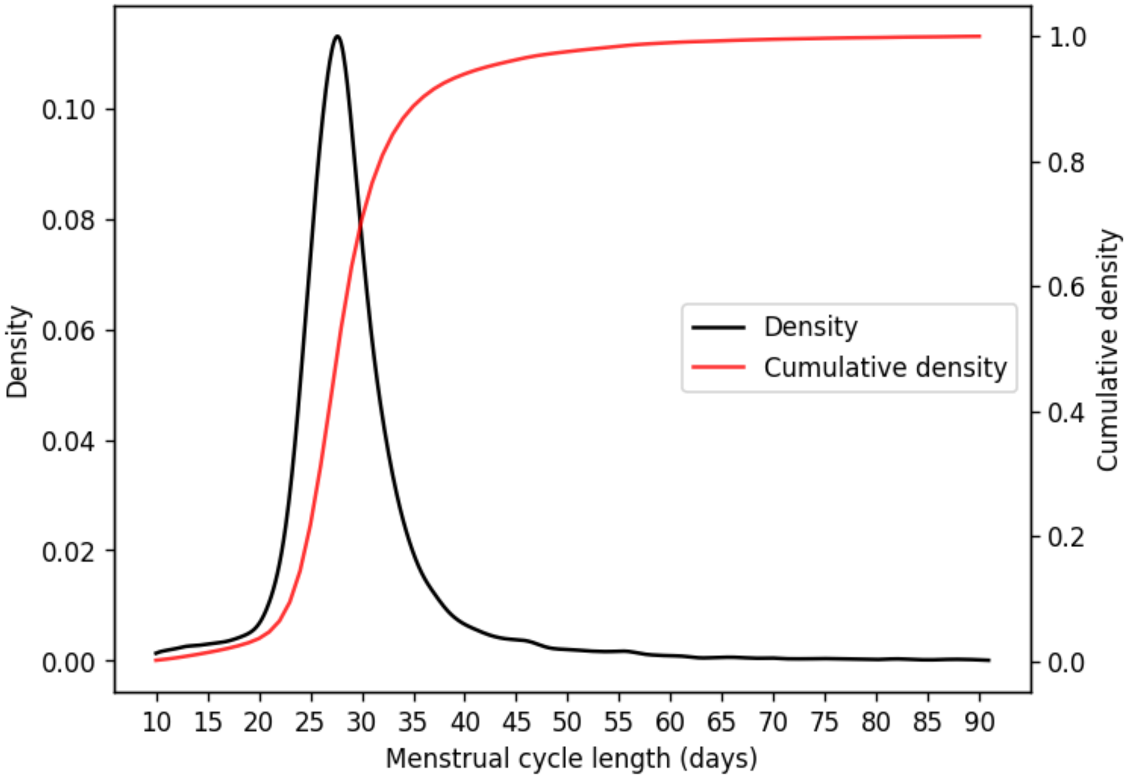


**Figure S2 By-participant distribution (density and cumulative density) of menstrual cycle length across 165,668 cycles from 12,608 participants in the AWHS**

Density and cumulative density were estimated by weighting the cycles with the inverse of the total number of cycles contributed by each participant.

Abbreviation: AWHS, Apple Women's Health Study.
